# Fractional versus standard BNT162b2 boosters after non-mRNA priming in Mongolia: 24-month immunogenicity and safety evidence from a randomised controlled trial

**DOI:** 10.1101/2025.09.04.25335126

**Authors:** Tsetsegsaikhan Batmunkh, Eleanor F G Neal, Otgonjargal Amraa, Nadia Mazarakis, Bolor Altangerel, Naranbaatar Avaa, Lkhagvagaram Batbayar, Khishigjargal Batsukh, Kathryn Bright, Tsogjargal Burentogtokh, Lien Anh Ha Do, Gantuya Dorj, John D Hart, Otgonbold Jamiyandorj, Khulan Javkhlantugs, Sarantsetseg Jigjidsuren, Frances Justice, Shuo Li, Khaliunaa Mashbaatar, Kerryn A Moore, Narantuya Namjil, Cattram Duong Nguyen, Batbayar Ochirbat, Unursaikhan Surenjav, Helen Thomson, Bilegtsaikhan Tsolmon, Paul V Licciardi, Claire von Mollendorf, Kim Mulholland

## Abstract

**Background:** Booster doses of COVID-19 vaccines help restore protection against waning immunity and emerging variants. Having found that fractional BNT162b2 boosters were non-inferior to standard boosters in eliciting anti-spike IgG responses at day 28 in Mongolian adults, we assessed long-term immunogenicity and safety in the same cohort.

**Methods:** In this randomised, controlled, non-inferiority trial, adults previously vaccinated with two doses of ChAdOx1-S, BBIBP-CorV, or Gam-COVID-Vac were randomly assigned (1:1) to receive a 15μg (fractional) or 30μg (standard) BNT162b2 booster. IgG concentrations and functional surrogate virus neutralisation test (sVNT) levels against Wuhan-Hu-1 and Omicron BA.1 were assessed over 24 months. A subset had IFN-γ cell-mediated-immunity (CMI) measured (Ag1/Ag2). Immune response GMRs (fractional:standard) were estimated from log-transformed values via multivariable linear regression. SARS-CoV-2 infections, adverse events, and serious adverse events (SAEs) were recorded. ClinicalTrials.gov Identifier: NCT05265065.

*Findings:* Of 601 participants enrolled between May 27 and Sept 30, 2022, 520 (86.5%) completed 24-month follow-up. Although IgG levels declined from six to 24 months, relative responses remained similar between arms at 18 months (GMR 1·08 [95% CI 0·97-1·22]) and 24 months (GMR 1·06 [95% CI 0·95-1·18]). In the CMI subset, IFN-γ responses peaked at day 28, waned to 18 months, and returned to baseline by 24 months, with fractional and standard arms similar at 24-month GMRs (Ag1 GMR 1·17 [95% CI 0·82–1·66]; Ag2 GMR 1·06 [95% CI 0·73–1·54]). Median sVNT inhibition against both Wuhan-Hu-1 and Omicron BA.1 was high and comparable between groups at 18 and 24 months (both 88% [95% CI 86-90]). SARS-CoV-2 infection was confirmed in 28 participants, with an additional 386 suspected infections after day 28, inferred from a ≥1·2-fold rise in IgG titres between study visits. No SARS-Cov-2 infections resulted in hospitalisation. Fifty-three SAEs were reported, evenly distributed between groups, with no vaccine-related events.

*Interpretation:* Fractional and standard BNT162b2 boosters showed comparable neutralising activity (sVNT) that persisted to 24 months, while binding IgG declined to baseline by 24 months. In the CMI subset, IFN-γ responses followed a similar trajectory, indicating alignment of humoral and cellular immunity over time. Despite widespread SARS-CoV-2 circulation, no COVID-19 hospitalisations or deaths occurred, and safety was reassuring. These data support fractional dosing as a pragmatic, cost-saving option.

**Funding:** Coalition for Epidemic Preparedness Innovations (CEPI). This study was supported by the Victorian Government’s Operational Infrastructure Support Programme.

**Panel: Research in Context:** *Evidence before this study:* We searched PubMed up to Aug 27, 2025, for English-language studies in adults assessing immune responses to fractional or reduced-dose COVID-19 boosters, focusing on mRNA vaccines and long-term follow-up. Search terms combined “fractional dose”, “low dose”, “reduced dose”, “booster”, “immunogenicity”, “antibodies”, “durability”, “persistence”, and related terms. Evidence is limited and largely confined to short- and medium-term follow-up. A trial in Thailand found half-dose AZD1222 after CoronaVac was non-inferior to a full dose at 14 and 90 days, with lower reactogenicity.^1^ A subsequent Thai study showed half-dose BNT162b2 or AZD1222 after CoronaVac achieved non-inferior immunogenicity at 28 and 90 days, particularly with longer intervals.^2^ In the UK, COV-BOOST reported similar anti-spike IgG responses with half- and full-dose BNT162b2 at three months,^3^ and by eight months, decay patterns varied by platform, but half-dose BNT162b2 tracked closely with full doses.^4^ In Belgium, the IMCOVAS trial showed low-dose mRNA-1273 and heterologous schedules were non-inferior to reference regimens up to one year, though intradermal vaccination was less effective.^5^ In Brazil, the FRACT-COV trial followed 1451 adults for six months: full-dose BNT162b2 elicited the highest titres, but fractional BNT162b2 outperformed full-dose AZD1222 and Sinovac.^6^ Across studies, fractional mRNA boosters generally induced robust short-term responses and, in some cases, non-inferior durability up to 12 months. However, no trial had reported a 24-month follow-up of fractional versus standard intramuscular BNT162b2 boosters.

*Added value of this study:* This randomised trial is the first to provide 24-month data comparing fractional (15 µg) and standard (30 µg) BNT162b2 boosters in adults primed with non-mRNA vaccines (ChAdOx1-S, BBIBP-CorV, Gam-COVID-Vac). Fractional dosing produced humoral and cellular responses equivalent to standard dosing: anti-spike IgG declined to baseline by 24 months, but neutralising activity and IFN-γ responses were preserved, with geometric mean ratios consistently close to unity. Age-stratified analyses showed higher early IgG responses in older adults that converged with younger participants by 18–24 months, indicating similar long-term durability across age groups. With 87% retention, detailed serological follow-up, and prespecified sensitivity analyses confirming that missingness did not bias results, this trial provides the first randomised long-term evidence from an LMIC setting. Our findings extend shorter-term results from Brazil, Thailand, and the UK by showing persistence of humoral and cellular immunity for two years.

*Implications of all the available evidence:* Fractional BNT162b2 boosters provide durable humoral and cellular immunity comparable to standard dosing, now shown up to 24 months. This supports dose-sparing as a feasible, cost-saving strategy to extend vaccine supply and improve equity, particularly in LMICs reliant on inactivated or adenoviral vector priming. While continued surveillance of emerging variants is needed, the available evidence indicates that fractional dosing can be used without compromising long-term protection.

## Introduction

The COVID-19 pandemic catalysed rapid global vaccination efforts, with booster doses recommended to increase the breadth and magnitude of protective immunity, especially to emerging variants. Optimising booster strategies is particularly important in low- and middle-income countries (LMICs), where constrained supply and limited access to updated vaccines remain a barrier. Fractional booster dosing, using reduced vaccine volumes, has been proposed as a dose-sparing approach to extend supply and reduce costs without compromising safety or immunogenicity.^7^

We conducted a randomised controlled phase 3 trial comparing a third dose fractional and standard booster of BNT162b2 (Pfizer–BioNTech) following primary vaccination with ChAdOx1-S (Oxford–AstraZeneca), BBIBP-CorV (Sinopharm), or Gam-COVID-Vac (Gamaleya) and previously reported 1 month immunogenicity and reactogenicity outcomes as well as 12-month follow-up data.^8,9^ These data showed comparable immunogenicity between dose groups for most priming strata, with durable IgG levels and stable neutralising responses to the ancestral strain and cross-reactivity to Omicron BA.1at 12 months post-vaccination.^8,9^ These findings are consistent with evidence from Brazil, Indonesia, Thailand, and the UK, albeit with shorter follow-up durations, supporting fractional dosing as a feasible and well-tolerated booster strategy.^1,4,10–12^ A phase IV trial in Brazil reported that although antibody titres were consistently higher following full-dose BNT162b2 boosters, fractional doses still elicited robust responses up to six months post-vaccination.^6^

Few studies have extended follow-up beyond one year, especially in LMIC populations primed with inactivated or viral vector vaccines. Longitudinal data are needed to assess antibody durability, long-term protection and safety, and to inform decisions about the need for ongoing booster doses, particularly as SARS-CoV-2 testing declines and antigenic mismatch with newer variants increases.^13–15^ While anti-spike IgG levels typically wane after boosting, several studies suggest responses plateau after six to 12 months, with durability varying by vaccine platform and schedule.^13,16,17^ Interpretation is further complicated by suspected SARS-CoV-2 infections, which may go undetected in the absence of anti-nucleocapsid testing, especially among recipients of inactivated vaccines.^18^ Recent Japanese studies have shown sustained immune memory up to 12 months after boosting with either BNT162b2 or a recombinant spike protein vaccine (S-268019-b), with broad neutralising responses and persistent memory B-cell and T-cell populations.^19,20^

This study reports 18- and 24-month follow-up immunogenicity data from our trial in Mongolia, providing the longest available data on fractional BNT162b2 boosters. We assess long-term antibody dynamics, intercurrent SARS-CoV-2 infections (documented and suspected), and adverse events. These data offer critical insight for policymaking in resource-limited settings and contribute to the evidence base on optimising COVID-19 booster strategies in the context of evolving variants, increasing population exposure, and hybrid immunity amidst constrained vaccine supply.

## Methods

### Trial design

This phase 3, double-blind, randomised, controlled, non-inferiority trial was conducted in Mongolia to evaluate the immunogenicity and safety of fractional versus standard BNT162b2 booster doses. The intervention compared a 15 μg (fractional) dose with the standard 30 μg dose of BNT162b2, administered as a third (booster) dose. Participants were followed for 24 months, with peripheral blood samples collected at baseline and at 28 days, six, 12, 18, and 24 months post-vaccination. The current analysis extends previous reports by including long-term outcomes at 18 and 24 months. The study was approved by the Mongolian Ministry of Health Ethics Committee (Decision #273, 05 April 2022) and the Royal Children’s Hospital Human Research Ethics Committee (HREC/81800/RCHM-2021) and conducted in accordance with the Declaration of Helsinki and Good Clinical Practice guidelines. The trial is registered with ClinicalTrials.gov (NCT05265065), and the protocol has been published previously.^8^

### Participants, randomisation, and blinding

Full trial methods have been described previously.^8^ Briefly, adults aged ≥18 years who had completed a two-dose primary vaccination series with BBIBP-CorV, ChAdOx1-S, or Gam-COVID-Vac were eligible. Participants were randomised (1:1) to receive a 15 μg (fractional) or 30 μg (standard) BNT162b2 booster dose, stratified by age group (18 – <50 years vs ≥50 years) and priming vaccine type. A subset of approximately 40% were included in a cell-mediated immunity (CMI) sub study, with age strata (18 – <50 years and ≥50 years) equally represented through a separate randomisation list. Due to laboratory processing capacity (10–20 samples/day), the first 10– 20 eligible participants per day were recruited into the CMI sub study, and daily recruitment ceased once the maximum number was reached.

### Procedures

Peripheral blood was collected at baseline and at 28 days, six, 12, 18, and 24 months post-booster. Anti-spike IgG was measured using the EUROIMMUN SARS-CoV-2 QuantiVac S1 IgG ELISA (EUROIMMUN Medizinische Labordiagnostika AG, Lübeck, Germany) and reported in binding antibody units (BAU)/mL, with titres classified as negative (<25·6 BAU/mL), borderline (25·6–35·2), or positive (>35·2). Cell-mediated immune responses were assessed using a QuantiFERON-based interferon-gamma (IFN-γ) release assay (IGRA) (QuantiFERON SARS-CoV-2 RUO; Qiagen, Hilden, Germany) in whole blood collected at baseline and at six, 12, and 24 months post-booster. Heparinised or EDTA-anticoagulated whole blood was stimulated with proprietary SARS-CoV-2 spike peptide pools (Ag1 and Ag2) and incubated for 16–24 hours, after which IFN-γ concentration was measured in IU/mL via enzyme-linked immunosorbent assay (ELISA), noting that anticoagulant type was not expected to influence IFN-γ readouts. Responses were considered positive if the IFN-γ level in antigen-stimulated wells exceeded the manufacturer’s threshold (e.g., ≥0·15 IU/mL above the nil control). Neutralising activity against Wuhan-Hu-1 and Omicron B.1.1.529 (BA.1) was assessed via the GenScript cPass surrogate virus neutralisation test (sVNT), with ≥30% inhibition considered positive.

Solicited adverse events (AEs) were monitored for three months post-booster, with unsolicited AEs and serious adverse events (SAEs) recorded throughout. Events were graded and classified using MedDRA terms. Data were collected and managed using REDCap.^21,22^

SARS-CoV-2 infections were tracked through participant self-report at enrolment and follow-up visits, including PCR or rapid antigen test (RAT) confirmation, symptoms, and illness severity. From March 2023 (month six onward), monthly SMS prompts were used to identify suspected infections. Participants reporting SARS-CoV-2 infection were invited to provide an acute phase nasopharyngeal swab for genomic sequencing and a convalescent blood sample 28 days later.

### Statistical Analysis

### Participant Recruitment and Follow-Up

Statistical methods used to describe recruitment procedures and baseline characteristics have been detailed previously.^8^ In this analysis, participant retention was summarised at each scheduled visit (28 days, six, 12, 18, and 24 months) using descriptive statistics. The number and percent of participants attending each visit, withdrawing, or lost to follow-up were summarised by study arm and primary vaccine.

### Assessment of Missingness

We systematically quantified missing data for immunogenicity endpoints, baseline characteristics, and other denominators within each analysis population and visit (baseline, day 28, six, 12, 18, and 24 months). In the main immunogenicity dataset, we calculated the number and percentage missing for anti-spike IgG concentrations and sVNT inhibition (Wuhan-Hu-1 and Omicron BA.1), overall and stratified by randomisation arm and priming vaccine. We also summarised completeness of key baseline covariates (age group, priming vaccine, vaccination dates) and the recorded study day of blood draw.

For the CMI sub study, we applied the same framework to interferon-γ responses (Ag1 and Ag2), tabulating the number and percentage missing at each visit, overall and by arm and priming strata. Because Ag1 and Ag2 had identical missingness, results are presented once for both antigens. To assess potential bias from loss to follow-up, we additionally summarised 24-month missingness of Ag1/Ag2 by baseline characteristics (age group, sex, prior infection, comorbidities, cigarette use, pregnancy), using cross-tabulations for categorical variables and medians (IQR) for continuous variables.

Analysis populations were pre-specified and included all randomised participants who received a study vaccine, categorised according to the dose received. Exclusions due to missing baseline or follow-up data were documented. Missing data rates were calculated both for the full enrolled cohort and within each analysis population to distinguish the impact of prior exclusions on data availability.

### Estimand Framework and Analysis Approach

As described previously, we used the Estimand Framework to ensure alignment between study objectives and statistical analysis.^23^ Estimand-to-analysis tables were prespecified in the statistical analysis plan, outlining the five estimand attributes for each endpoint.^8,9^ For long-term immunogenicity, the primary estimands were the geometric mean ratio (GMR) of IgG and the median surrogate virus neutralisation levels at six, 12, 18, and 24 months after a single fractional (15 µg) versus standard (30 µg) dose of BNT, in adults aged ≥18 years in Mongolia, regardless of SARS-CoV-2 infection or receipt of a fourth dose. Post-boost SARS-CoV-2 infection was treated as an intercurrent event using the Treatment Policy strategy. Supplementary analyses using a Hypothetical Strategy (i.e., setting infections occurring >14 days post-boost to “no infection”) were prespecified, but were not implemented for two reasons. Firstly, the number of adjudicated PCR-confirmed cases was small (n=28) and balanced between arms; and secondly, infection ascertainment was incomplete, with longitudinal serology indicating substantial undocumented exposure, creating a non-ignorable risk of misclassification of the intercurrent event and rendering the hypothetical estimand difficult to interpret. Given these considerations, the Treatment Policy strategy was retained for primary inference as the most robust reflection of real-world conditions. Primary analyses were conducted on complete cases. As prespecified in the statistical analysis plan, if missingness in the primary outcome (anti-spike IgG) exceeded 10%, sensitivity analyses using multiple imputation by chained equations (MICE) were performed for GMCs by arm and GMRs at Day 28, six, 12, 18 and 24 months, with estimates combined across imputations using Rubin’s rules.^24,25^

### Long-term Anti-Spike IgG Levels

IgG levels were summarised by visit, arm, priming stratum, and age groups using geometric means and 95% CIs. GMRs were estimated using linear regression on log-transformed anti-spike IgG levels, adjusted for age group, priming strata, timing of blood draw, dosing intervals, and baseline IgG. Day 28 endpoints were previously analysed under a non-inferiority framework, while later time points (six, 12, 18, and 24-month outcomes) were analysed as continuous variables under a superiority framework, with effect sizes and 95% confidence intervals (CIs). Interaction terms allowed estimation of GMRs within priming strata. GMRs were not estimated separately within age groups, as the trial was not designed for such subgroup analysis. To avoid over-interpretation of post-hoc comparisons, we instead present descriptive GMCs by age group to illustrate overall trends in humoral immunity.

### Long-term Cell Mediated Immune Responses (QuantiFERON interferon-gamma release assay IGRA)

Geometric means and 95% CIs of IFN-γ concentrations were summarised by visit, study arm, and priming stratum; between-group comparisons are presented as GMRs with 95% CIs. IFN-γ concentrations were log-transformed and modelled using linear regression to estimate GMRs at baseline, day 28, six, 12, 18, and 24 months post-booster. At baseline, models were adjusted for age group, priming vaccine, days between doses 1–2, days between dose 2 and the study dose, and the study day of blood draw. At post-baseline visits (day 28 onward), models were adjusted for the same variables plus baseline IFN-γ. Responses are reported separately for Ag1 and Ag2 peptide pools and are presented numerically and graphically. As with the humoral analyses, age-stratified results are presented descriptively.

### Long-term sVNT Inhibition Responses to Wuhan-Hu-1 and Omicron BA.1

To assess the durability of neutralising responses, median and IQR of sVNT percent inhibition against Wuhan-Hu-1 and Omicron BA.1 were calculated at each time point, stratified by study arm and priming strata. Results are presented numerically and graphically.

### Documented and Suspected Intercurrent SARS-CoV-2 Infections

To visualise SARS-CoV-2 infections during follow-up, an epidemiological curve was generated showing documented infections by study week. To explore potential undocumented infections, individual-level line plots of anti-spike IgG titres were generated by study arm and priming strata, with each line representing individual participant’s titres at baseline, day 28, six months, 12 months, 18 months, and 24 months. Undocumented infections were defined as a ≥1·2-fold increase in IgG levels between consecutive visits without a reported SARS-CoV-2 infection.^26^ Documented and suspected undocumented infections were tabulated, and differences between study arms and priming strata were assessed using the chi-squared test (α<0·05).

### Adverse and Serious Adverse Events

AEs and SAEs were summarised by severity, causality, and MedDRA System Organ Class (SOC). Causality was assessed independently by both the investigator and the study sponsor. Events occurring within the 24-month follow-up window (defined as <787 days post-randomisation) were included. Descriptive statistics (counts and percentages) were reported. All statistical analyses were conducted using Stata version 18.5.^27^

## Results

### Participant recruitment and follow-up

Baseline characteristics have been described previously for the 601 enrolled participants.^8^ At 18 months, 529 participants were analysed (265 in the fractional-dose arm and 264 in the standard-dose arm; Figure 1). At 24 months, 520 participants were analysed (260 in each arm). No participant reported receiving a fourth COVID-19 vaccine dose in the community.

**Figure 1.**
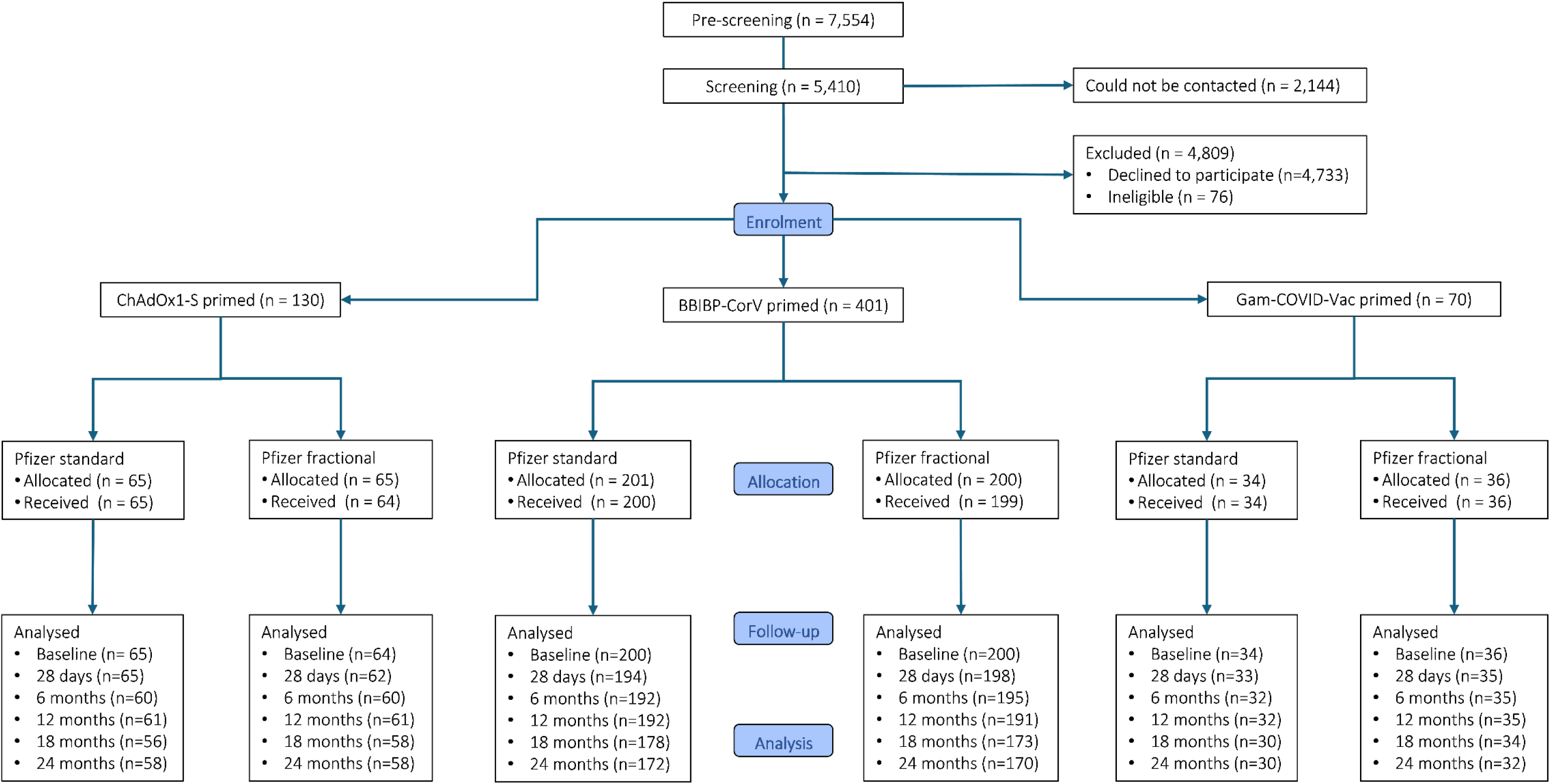
Trial flow-chart stratified by priming vaccine (ChAdOx1-S, BBIBP-CorV, and Gam-COVID-Vac) and study arm (fractional [15μg] or standard [30μg] BNT162b2 booster)

By 24 months, 59 (9·8%) participants had withdrawn; 35 due to relocation, 17 due to voluntary withdrawal, and six due to death (four in the standard dose arm (three primed with BBIBP-CorV and one with Gam-COVID-Vac) and two in the fractional dose arm (both BBIBP-CorV primed). All deaths were unrelated to study vaccine. One participant was withdrawn at the investigator’s discretion due to a pre-existing medical condition. A further 22 (3·7%) participants were lost to follow-up, leading to a total loss of 13·5%.

Of the total cohort, 256 participants (42·6%) were enrolled in the CMI sub study, with equal representation from the standard-dose arm (128/301, 42·5%) and the fractional-dose arm (128/300, 42·7%). Evaluable QuantiFERON IGRA samples were obtained at baseline (242/256, 94·5%), day 28 (238/256, 93·0%), six months (242/256, 94·5%), 12 months (241/256, 94·1%), 18 months (220/256, 85·9%), and 24 months (222/256, 86·7%). At 24 months, evaluable samples were balanced by arm, with 85·2% (109/128) in the fractional arm and 88·3% (113/128) in the standard arm.

## Immunological responses up to 24 months post-booster

### Anti-Spike IgG levels

Missingness for immunological outcomes and covariates was low at early timepoints and increased at later visits, with 13·5% of participants missing anti-spike IgG and sVNT data at 24 months due to withdrawal or loss to follow-up (Supplementary Table 1). At 24 months, missingness of immune response data was higher among participants aged 18 – <50 years than those aged ≥50 years, and among males, but rates were similar across treatment groups and priming vaccines, with no strong evidence of systematic attrition (Supplementary Table 2).

Anti-spike IgG levels declined from day 28 to six months and remained stable through 12 months.^8,28^ At 18 and 24 months, levels continued to decline, and by 24 months had returned to baseline concentrations, with comparable responses between fractional and standard dose groups (18 months GMR 1·08 [95% CI 0·97–1·21]; 24 months: 1·06 [95% CI 0·95–1·18]; (Supplementary Table 3). Patterns were similar across priming vaccine groups, with no consistent differences between arms (Figure 2).

**Figure 2.**
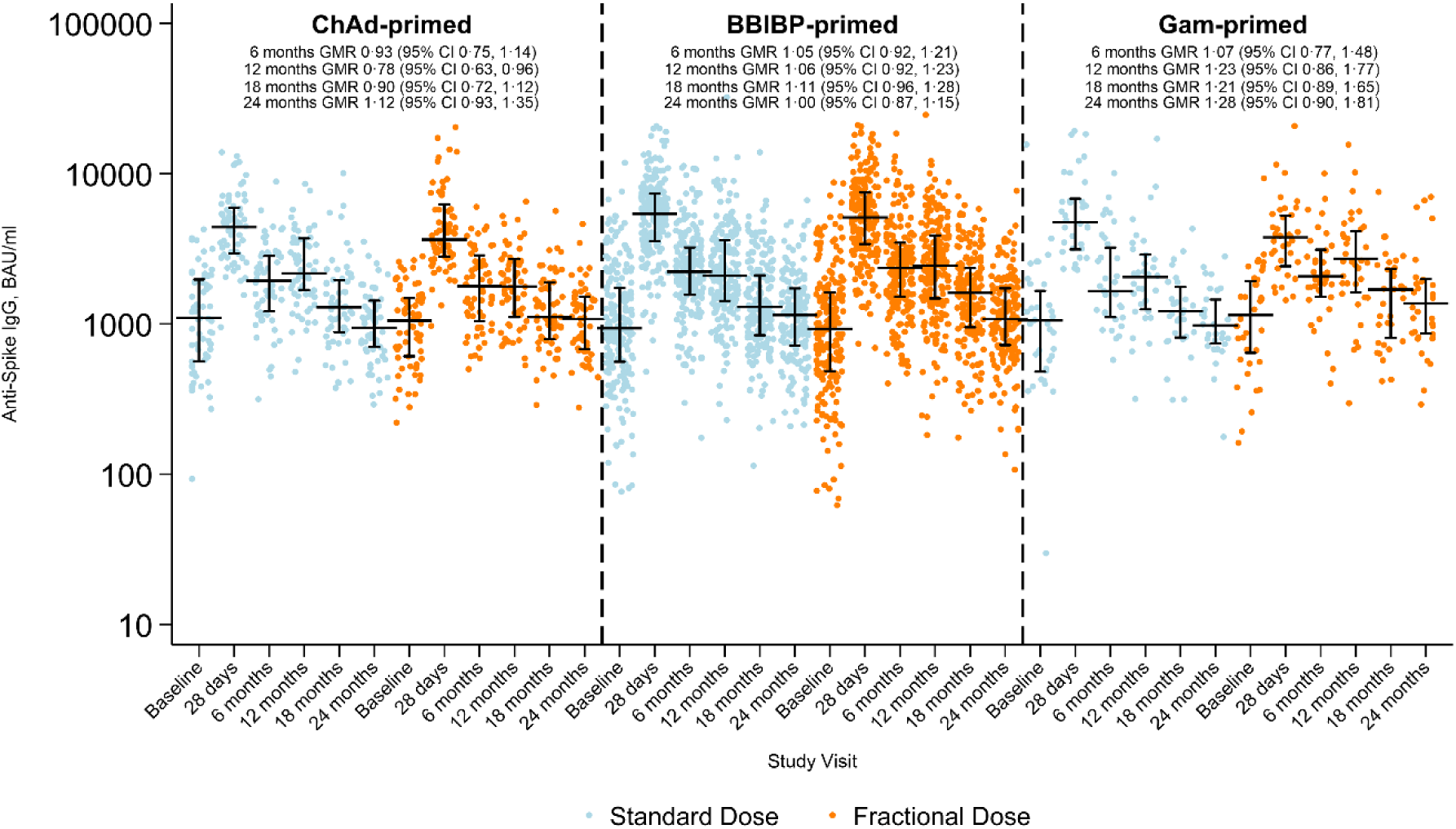
Anti-spike IgG by study visit, priming vaccine, and study arm. Horizontal black lines with vertical bars indicate the median and Interquartile range. GMR: Geometric mean ratio at six, 12, 18, and 24 months for levels between fractional and standard dose arms, adjusted for age group, priming vaccine, duration between first and second dose, duration between second and third (study) dose, study day of blood draw, and baseline anti-spike IgG

Sensitivity analyses using multiple imputation yielded IgG GMC and GMR estimates at Day 28, six, 12, 18 and 24 months that were comparable with the primary complete-case analyses, with no change in inference (Supplementary Table 4).

Across both study arms, baseline anti-spike IgG GMCs were higher among participants aged ≥50 years compared with those aged 18 – <50 years (Supplementary Table 5). Following booster vaccination, peak responses at day 28 were robust in both age groups, with similar GMCs between standard- and fractional-dose recipients. IgG levels declined thereafter but were sustained above baseline throughout follow-up. At six and 12 months, GMCs remained slightly higher in older participants, whereas by 18 and 24 months, levels had converged between age groups. By 24 months, GMCs were approximately 1,000–1,200 BAU/mL across arms and age strata, indicating durable responses with no differences between dosing groups.

### Long-term Cell Mediated Immune Responses: Interferon-γ (Ag1 and Ag2)

Within the CMI sub study (n = 256), data completeness for interferon-γ concentrations was high at early visits (>90% through 12 months). Missingness increased modestly at later visits, reaching 14·1% (36/256) at 18 months and 13·3% (34/256) at 24 months, with similar rates across study arms, priming strata, sex, and age groups. Full breakdowns are provided in Supplementary Tables 6 and 7.

IFN-γ responses peaked at day 28, waned to 18 months, and showed a modest rise by 24 months, mirroring the IgG kinetics (Supplementary Tables 8 and 9). Between-arm differences (fractional vs standard) were small at every visit. For Ag1, adjusted GMRs were 1·20 (0·83–1·73) at baseline, 1·19 (0·91–1·56) at day 28, 1·06 (0·77– 1·47) at six months, 0·98 (0·71–1·37) at 12 months, 1·18 (0·78–1·78) at 18 months, and 1·17 (0·82–1·66) at 24 months. For Ag2, GMRs were 1·18 (0·83–1·69) at baseline, 1·06 (0·81–1·38) at day 28, 1·14 (0·82–1·59) at six months, 1·14 (0·84–1·54) at 12 months, 0·99 (0·70–1·40) at 18 months, and 1·06 (0·73–1·54) at 24 months. All 95% CIs included 1, indicating little difference between dosing strategies. Patterns were consistent across priming strata; the only near-signal was lower Ag1 in the fractional arm at 12 months among Gam-COVID-Vac (GMR 0·56, 95% CI 0·30–1·04), based on small numbers. IFN-γ responses for Ag1 and Ag2 are shown in Figure 3.

**Figure 3.**
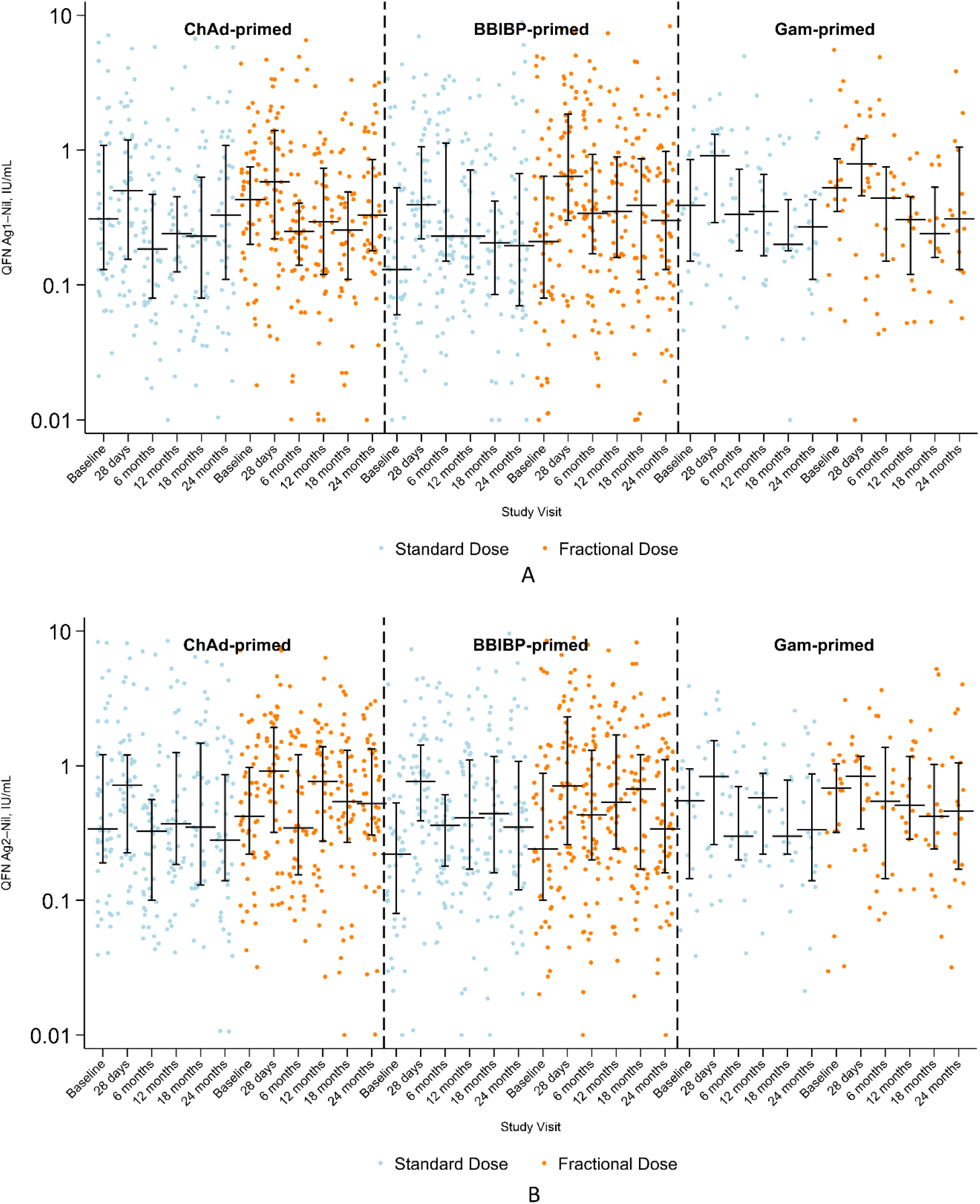
IFN-γ concentrations for Ag 1 (A) and Ag 2 (B) by study visit, priming vaccine, and study arm. Horizontal black lines with vertical bars indicate the median and interquartile range. GMR: geometric mean ratio at six, 12, 18, and 24 months for levels between fractional- and standard-dose arms, adjusted for age group, priming vaccine, duration between first and second dose, duration between second and third (study) dose, study day of blood draw, and baseline IFN-γ concentration for the respective antigen.

Supplementary Table 10 presents geometric mean IFN-γ concentrations (Ag1 and Ag2) with 95% CIs by study arm, stratified by age group (18 – <50 years vs ≥50 years) across all study visits. IFN-γ responses rose after the booster in both age groups, peaking at day 28 and waning by six to 12 months. From 18 months onward, concentrations were generally higher in participants ≥50 years compared with those 18 – <50 years, particularly for Ag2. Fractional-dose recipients frequently showed responses comparable to or greater than standard-dose recipients across age groups. By 24 months, both age groups retained low but detectable responses, with no evidence that fractional dosing impaired durability.

### Wuhan-Hu-1 SARS-CoV-2 sVNT inhibition

Median RBD–hACE2 inhibition was similar between study arms through the first 12 months and remained high following booster vaccination (Supplementary Table 11 and Figure 4). Previously published results showed stable median inhibition from baseline to day 28, with an increase by six months that was sustained at 12 months across both study arms and all priming schedules.^8,28^ At 18 months, median inhibition was 88% (IQR 87–90) in the standard-dose arm and 88% (86–90) in the fractional-dose arm. At 24 months, values remained stable and sustained above baseline levels, at 88% (86–89) and 88% (86–90), in the standard and fractional dose arms, respectively. This pattern was also observed for Omicron BA.1. Patterns were similar across priming schedules, with median inhibition sustained at high levels through 24 months in both arms. These findings indicate durable neutralising activity to 24 months, contrasting with the waning of binding IgG.

**Figure 4.**
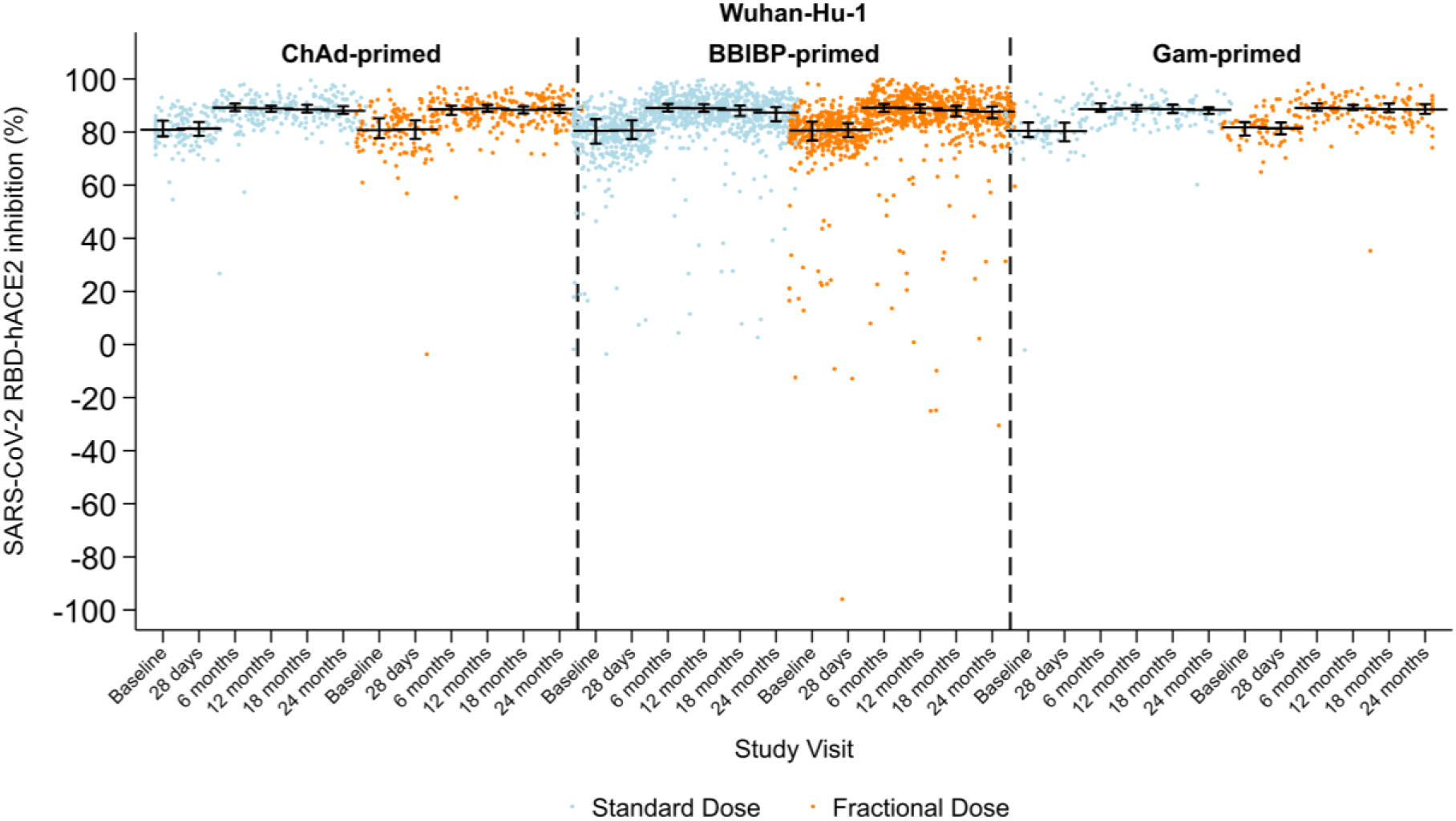
SARS-CoV-2 sVNT inhibition (%) against Wuhan-Hu-1 by study visit, priming vaccine, and study arm. Horizontal black lines with vertical bars indicate the median and interquartile range.

### Omicron BA.1 SARS-CoV-2 sVNT inhibition

Previously published data showed that sVNT inhibition against BA.1 increased markedly by day 28 and was sustained through 12 months, with minimal differences between standard and fractional dosing schedules (Supplementary Table 12 and Figure 5.^8,28^ Median inhibition peaked at day 28 (82% in the standard arm and 81% in the fractional arm), declined slightly by six months, and remained stable at 12 months across all priming groups.

**Figure 5.**
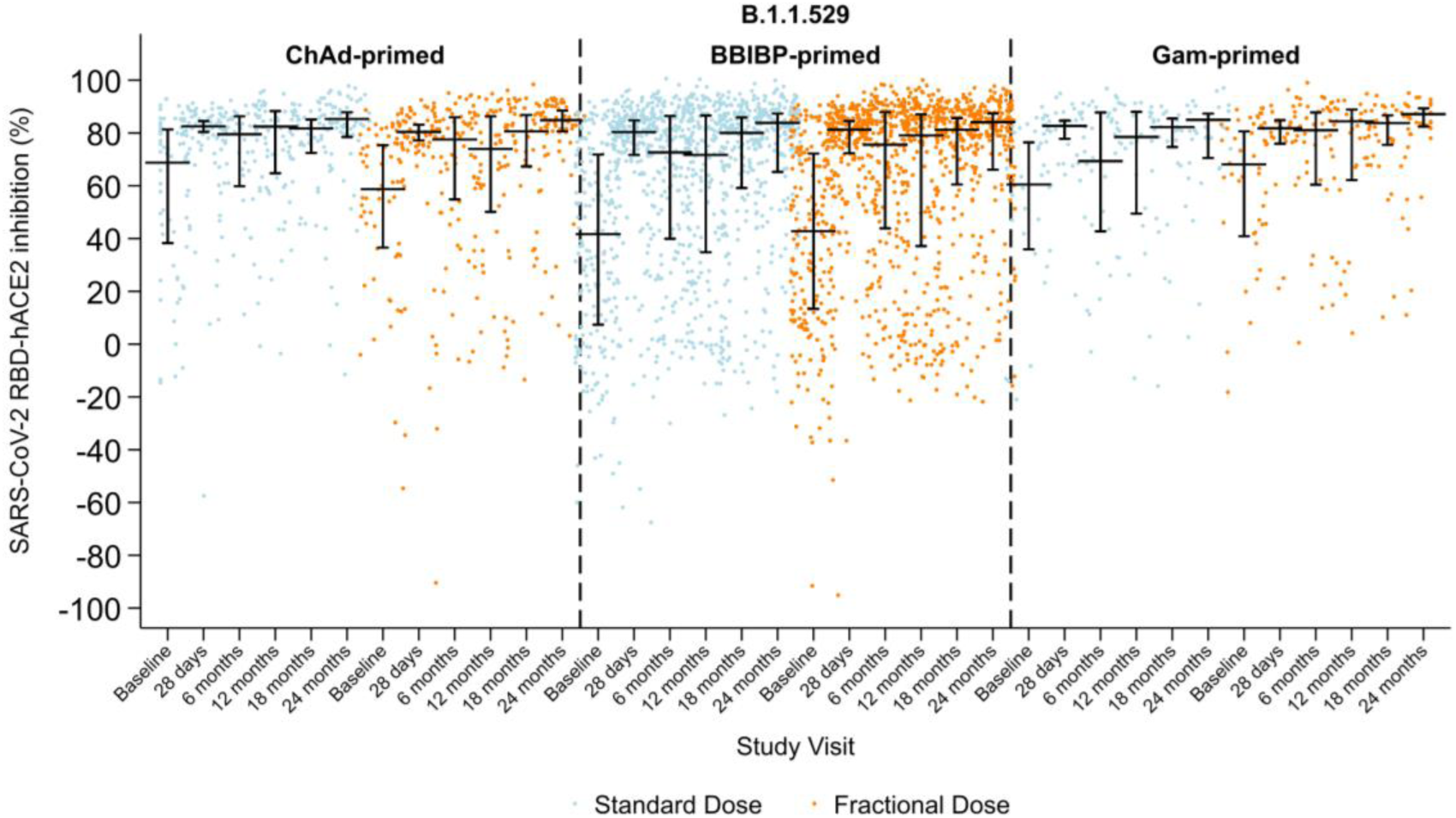
SARS-CoV-2 sVNT inhibition (%) against Omicron BA.1 by study visit, priming vaccine, and study arm. Horizontal black lines with vertical bars indicate the median and interquartile range.

At 18 months, median inhibition remained high: 81% (IQR 61–86) in the standard-dose arm and 81% (64–86) in the fractional-dose arm. By 24 months, inhibition had increased modestly to 84% (71–88) in the standard arm and 85% (70–88) in the fractional arm. Neutralising responses at 24 months were comparable across priming schedules, with inhibition levels increasing or remaining stable in both arms. Importantly, BA.1 neutralisation was modest at baseline (median ∼52%) but rose after boosting and remained high at 24 months (∼84–85%), in contrast to binding IgG which had waned back to baseline.

### Documented and Suspected Intercurrent SARS-CoV-2 Infections

Changes in individual anti-spike IgG levels between study visits, stratified by study arm and priming vaccine, are shown in Figure 6. A fold increase of ≥1·2 between timepoints is considered indicative of a suspected SARS-CoV-2 infection.^26^ Overall, antibody kinetics appeared similar between fractional and standard dose groups over time.

**Figure 6.**
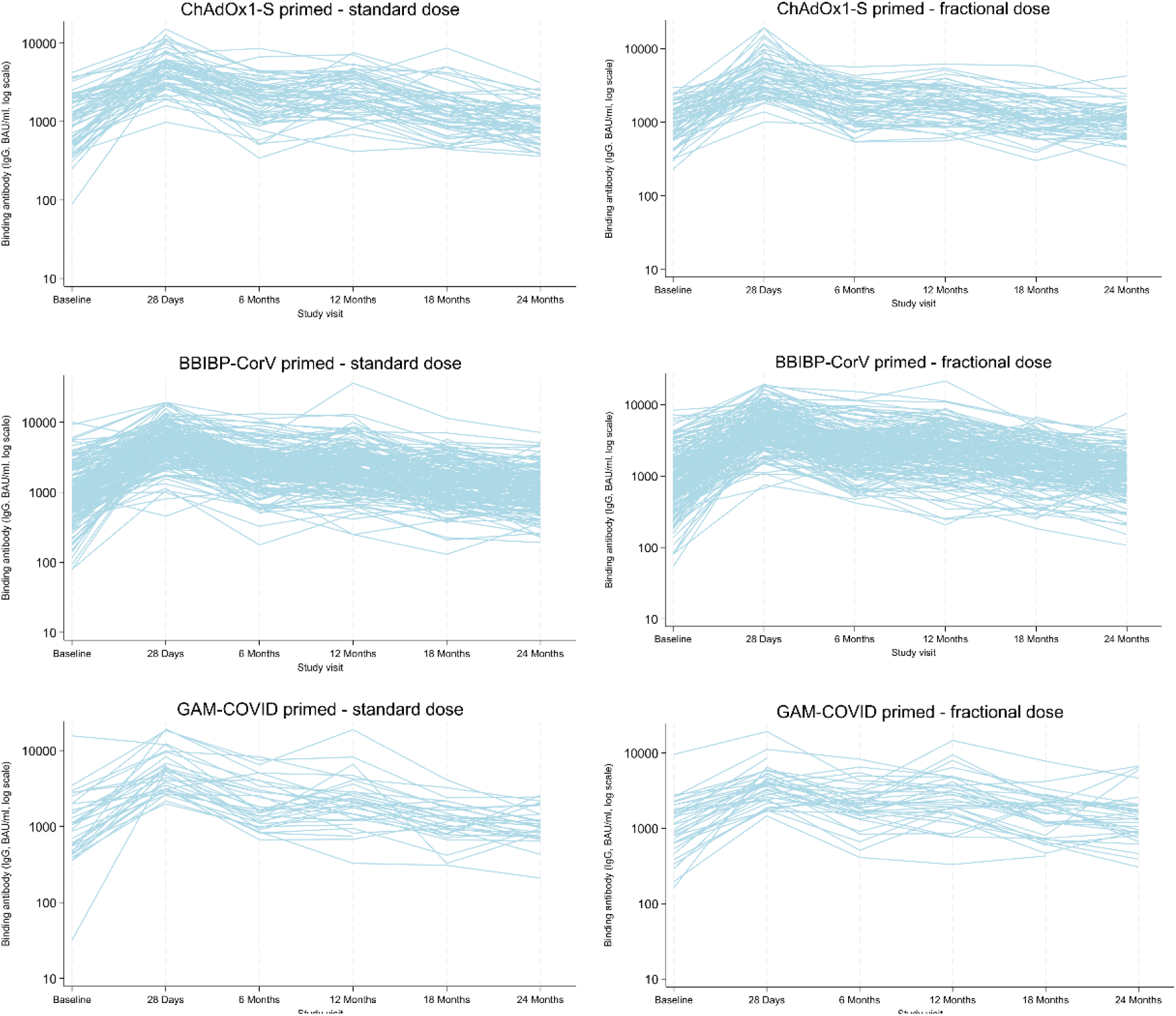
Change in individual binding antibody titres between Baseline and 28 days, 28 days and six months, six and 12 months, 12 and 18 months, and 18 and 24 months post-booster, by priming vaccine and study arm

Table 1 summarises numbers (%) of participants with fold changes ≥1·2 in anti-spike IgG levels without a documented SARS-CoV-2 infection and documented SARS-CoV-2 infections across study visits, stratified by study arm and priming strata. Documented infections remained rare across the study period (Supplementary Figure 1), with a cumulative incidence of 4·7% (28/601) by 24 months and no meaningful differences between study arms (14/301 (4·7%) standard arm and 14/300 (4·7%) fractional arm). The majority of documented SARS-CoV-2 infections occurred in participants aged 18 – <50 years (23/28 (82·1%), with comparatively few in those aged ≥50 years (5/28 (17·9%). As previously reported, most increases in anti-spike IgG levels within 28 days reflected the recent booster dose, and suspected infections were more common between six and 12 months than at earlier timepoints.

**Table 1.** Documented and suspected SARS-CoV-2 infections between study visits, stratified by study arm and priming vaccine^a^.

| Priming strata | Fold change $\geq 1.2$ in anti-spike IgG levels without documented SARS-CoV-2 infection n/N<sup>b</sup> (%) | | Documented SARS-CoV-2 infections, n/N<sup>c</sup> (%) | |
| --- | --- | --- | --- | --- |
|  | Standard dose | Fractional dose | Standard dose | Fractional dose |
| **Baseline - day 28<sup>d</sup>** |  |  |  |  |
| All | 282/291 (96.9) | 285/294 (96.9) | 1/292 (0.3)<sup>e</sup> | 1/295 (0.3)<sup>f</sup> |
| ChAdOx1-S | 63/64 (98.4) | 61/62 (98.4) | 1/65 (1.5) | 0/62 (0.0) |
| BBIBP-CorV | 187/194 (96.4) | 189/197 (95.9) | 0/194 (0.0) | 1/198 (0.5) |
| Gam-COVID-Vac | 32/33 (97.0) | 35/35 (100.0) | 0/33 (0.0) | 0/35 (0.0) |
| **Day 28 - six months** |  |  |  |  |
| All | 8/271 (3.0) | 20/280 (7.1) | 8/279 (2.9) | 8/288 (2.8) |
| ChAdOx1-S | 2/56 (3.6) | 2/58 (3.4) | 4/60 (6.7) | 2/60 (3.3) |
| BBIBP-CorV | 5/184 (2.8) | 12/189 (6.4) | 4/188 (2.1) | 5/194 (2.6) |
| Gam-COVID-Vac | 1/31 (3.2) | 6/33 (18.2) | 0/31 (0.0) | 1/34 (2.9) |
| **Six - 12 months** |  |  |  |  |
| All | 107/275 (38.9) | 92/279 (33.0) | 4/279 (1.4) | 3/282 (1.1) |
| ChAdOx1-S | 27/57 (47.4) | 17/57 (29.8) | 2/59 (3.4) | 1/58 (1.7) |
| BBIBP-CorV | 69/186 (37.1) | 59/187 (31.6) | 2/188 (1.1) | 2/189 (1.1) |
| Gam-COVID-Vac | 11/32 (34.4) | 16/35 (45.7) | 0/32 (0.0) | 0/35 (0.0) |
| **12 - 18 months** |  |  |  |  |
| All | 34/261 (13.0) | 38/264 (14.4) | 0/261 (0.4) | 0/264 (0.0) |
| ChAdOx1-S | 6/56 (10.7) | 7/57 (12.3) | 0/56 (1.8) | 0/57 (0.0) |
| BBIBP-CorV | 26/175 (14.9) | 28/173 (16.2) | 0/175 (0.0) | 0/173 (0.0) |
| Gam-COVID-Vac | 2/30 (6.7) | 3/34 (8.8) | 0/30 (0.0) | 0/34 (0.0) |
| **18 - 24 months** |  |  |  |  |
| All | 45/252 (17.9) | 42/251 (16.7) | 1/253 (0.4) | 2/252 (0.4) |
| ChAdOx1-S | 9/55 (16.1) | 13/56 (23.2) | 1/56 (1.8) | 1/57 (1.8) |
| BBIBP-CorV | 30/168 (17.9) | 21/163 (12.9) | 0/168 (0.0) | 1/163 (0.6) |
| Gam-COVID-Vac | 6/29 (20.7) | 8/32 (25.0) | 0/29 (0.0) | 0/32 (0.0) |

Abbreviations: IgG – immunoglobulin G; Footnotes: <sup>a</sup> Suspected SARS-CoV-2 infections were defined as a $\geq 1.2$ -fold increase in anti-spike IgG levels between consecutive study visits without a documented SARS-CoV-2 infection; <sup>b</sup> Documented SARS-CoV-2 infection excluded from numerator and denominator; <sup>c</sup> Number who attended both baseline and day 28 visits, day 28 and six-month visits, six and 12 month visits, 12 and 18 month visits, and 18 and 24 month visits; <sup>d</sup> Documented infections were only considered valid if they occurred between 14 and 28 days but are listed for completeness, and IgG levels were expected to rise at 0–28 days due to the timing of the measurements shortly after booster administration; <sup>e</sup> One participant in the standard-dose arm (ChAdOx1-S–primed) had a documented SARS-CoV-2 infection 4 days after booster; <sup>f</sup> One participant in the fractional-dose arm (BBIBP-CorV–primed) had a documented SARS-CoV-2 infection 18 days after booster.

Updated findings from 12 to 24 months showed no documented infections between 12 and 18 months, while suspected infections occurred in 13·0% (34/261) of the standard-dose group and 14·4% (38/264) of the fractional-dose group. Between 18 and 24 months, suspected infection rates were 17·9% (45/252) in the standard-dose group and 16·7% (42/251) in the fractional-dose group, with documented infections remaining low (0·4% [1/253] vs 0·8% (2/252).

### Adverse and Serious Adverse Events

From baseline to 24 months post-booster, 76 AEs and 53 SAEs were recorded (Supplementary Tables 13 and 14, and Supplementary Figure 2). The distribution of AEs remained balanced across study arms, with 38 (50·0%) occurring in each of the fractional and standard groups. Most AEs (48/76, 63·2%) were classified as mild, and the majority (49/76, 64·5%) were deemed unrelated to the study vaccine. A smaller proportion (27/76, 35·5%) were assessed as possibly or probably vaccine-related, including cases such as irregular menstruation, furuncles, headache, and hypertension, typically occurring within the first 90 days post-booster. Nearly all AEs (68/76, 89·5%) resolved completely, with a median duration of 10·5 days (IQR 5·5 – 28·5 days); among events resolving with sequelae (8/76,10·5%), the median duration was 18 days (IQR 9·5 – 41 days).

Up to 24 months 53 SAEs were reported, including a range of acute and chronic medical conditions with onset times extending up to 690 days post-vaccination. The majority were moderate (19/53, 36·0%) or severe (26/53, 49·1%) in intensity. Two events (3·8%) were considered potentially life-threatening, and six SAEs (11·3%) resulted in death. Fatal cases included sudden suicide (n=1), gastric cancer (n=1) cardiac events (n=1), sudden death (n=2), and decompensated diabetes (n = 1), all determined to be unrelated to the study vaccine. Of the 53 SAEs, 26 (49·1%) occurred in the fractional dose group and 27 (50·9%) in the standard dose group. Across both arms, 24/53 (45·3%) of SAEs resolved with sequelae, and no vaccine-related SAEs were identified.

## Discussion

This 24-month follow-up of a randomised trial in Mongolia provides the longest prospective comparison of fractional (15 µg) and standard (30 µg) BNT162b2 boosters after priming with non-mRNA vaccines (ChAdOx1-S, BBIBP-CorV, Gam-COVID-Vac). Binding IgG rose post-boost and waned towards baseline by 24 months without material differences between dose arms. In a prespecified subset, IFN-γ responses to Ag1/Ag2 showed the same trajectory, peaking at day 28, waning thereafter, and returning to baseline by 24 months, indicating alignment of humoral and cellular immunity over time. Neutralising activity against both ancestral (Wuhan-Hu-1) and Omicron BA.1 variants remained high through 24 months in both arms, indicating consistent cross-protective responses across strains. Documented SARS-CoV-2 infections were uncommon, but serology suggested under-ascertainment of largely asymptomatic exposure, with similar rates of suspected though undocumented infection across study arms. The high prevalence of asymptomatic or mildly symptomatic infections among vaccinated individuals makes self-reported status an unreliable metric, reinforcing the importance of serological surveillance in monitoring post-booster exposure. Serious adverse events were uncommon and balanced between groups. Taken together, these findings support fractional BNT162b2 dosing as a pragmatic, cost-saving option for booster programmes, particularly in resource-constrained settings.

Binding IgG rose substantially after boosting, remained stable between six and 12 months, and approached baseline by 24 months, with no differences between dose groups. Early arm-specific differences (lower 12-month titres in ChAdOx1-S–primed fractional recipients and a reversal among Gam-COVID-Vac–primed participants with higher titres in the fractional arm from 12 months) were not sustained, indicating transient kinetics rather than durable divergence by dose. Although age-stratified analyses were exploratory, baseline and day 28 IgG levels were higher in those ≥50 years, but titres converged with those of younger participants by 18 to 24 months, suggesting broadly similar long-term humoral durability across age groups. Thus, the long-term durability signal in our cohort reflects neutralising activity (sVNT) rather than binding IgG. Our findings extend those of shorter-term studies, including two Brazilian phase IV trials, where fractional BNT162b2 boosters induced lower titres than full doses but outperformed AZD1222 and Sinovac through 182 days of follow-up.^6,12^ In Japan, 12-month studies reported durable responses to BNT162b2 and the recombinant spike protein vaccine S-268019-b, with broad neutralisation and preserved immune memory.^19,20^ The COVE trial also demonstrated sustained responses to mRNA-1273 boosters over 24 months, particularly with longer prime-boost intervals.^29^ Modelling studies have suggested slower IgG decay following low-dose mRNA boosters (25 µg vs 100 µg);^30^ however, in our study using a 15 µg fractional BNT162b2 booster (vs standard 30 µg), decay rates were similar between fractional and standard dose arms, further supporting fractional dosing as a feasible long-term strategy.

Cell-mediated immunity persisted to 24 months in both arms. Ag1 captures CD4⁺ epitopes, whereas Ag2 captures both CD4⁺ and CD8⁺ epitopes, the latter providing a broader reflection of infection-relevant T-cell responses. IFN-γ responses to Ag1 and Ag2 rose post-booster, declined to about 12 months, and then stabilised, with no consistent differences by dose or priming regimen. Concordant Ag1/Ag2 trajectories indicate preserved T-cell memory breadth, which, given the relative conservation of T-cell epitopes across variants, plausibly contributes to protection from severe outcomes despite waning binding IgG.^31–33^ In exploratory analyses, IFN-γ responses, particularly to Ag2, were relatively well preserved in adults ≥50 years from 18 months onward, consistent with sustained T-cell memory. These cellular data complement the neutralising activity signal and are consistent with the absence of COVID-19–related hospitalisations or deaths in our cohort.

Neutralising antibody responses, measured by sVNT inhibition, remained high and stable through 24 months for both Wuhan-Hu-1 and Omicron BA.1, with minimal decline from 12 to 24 months, and no material differences by dose or priming strata. Within the BBIBP-CorV–primed cohort, sVNT inhibition values showed greater variability for both Wuhan-Hu-1 and Omicron BA.1 than in the ChAdOx1-S or Gam-COVID-Vac cohorts, despite similar medians, suggesting heterogeneous priming and/or undocumented interim exposure. We did not power the study for formal variance comparisons, so this observation should be interpreted cautiously. Nonetheless, these findings suggest preserved functional antibody activity and cross-neutralisation to Omicron BA.1, despite receipt of ancestral-strain vaccines. Although Omicron BA.1 binding titres were not assayed here, prior data indicate binding responses to Omicron are typically lower than those to ancestral strains, while sVNT activity is relatively preserved.^8,17^ Emerging evidence suggests substantially lower neutralisation of newer lineages (e.g., XBB and JN.1), even after bivalent boosting, underscoring the need to align future antigen panels with circulating variants.^15^ Against this backdrop, the persistence of IFN-γ responses in our CMI subset, together with the relative conservation of T-cell epitopes across SARS-CoV-2 variants, supports sustained protection against severe disease even as serum neutralisation drifts, underscoring the importance of aligning booster timing with local epidemiology and variant evolution.^15,31–33^

During follow-up, only 28 PCR-confirmed SARS-CoV-2 infections were reported, distributed evenly between arms, with most cases in participants aged 18 – <50 years, possibly reflecting greater community exposure and mobility, rather than differences in vaccine-derived protection. Longitudinal serology indicated considerable under-ascertainment. Many participants, particularly those primed with Gam-COVID-Vac and those in the fractional arm, had ≥1·2-fold rises in anti-spike IgG without a recorded test, consistent with asymptomatic or untested exposure. These episodes clustered at six to 12 months post-boost, declined thereafter, and rose slightly by 24 months, suggesting ongoing low-level transmission. Similar trends and under-detection were observed in the REDUCE cohort study in Austria, which recorded 1,672 PCR-confirmed SARS-CoV-2 infections over 24 months among 3,859 individuals who had received three 30 µg doses of BNT162B2.^34^ As case identification relied on national PCR testing databases, where testing was largely symptom-driven, asymptomatic infections were likely under-detected. In that setting, vaccine effectiveness against infection declined from 81·6% (95% CI 80·0%–83·2%) during the first nine months to 38·2% (95% CI 35·8%–40·6%) between months 10 and 24, underscoring the impact of waning protection and persistent viral circulation even in well-vaccinated populations.^34^ While REDUCE used PCR-confirmed endpoints and our study captured both documented and likely undocumented exposures through serological inference, both studies illustrate the growing challenges of infection surveillance in the post-acute pandemic phase.

Over the 24 month period post-vaccination, the safety profile remained favourable. Most adverse events were mild or moderate and occurred more frequently in the standard-dose group, but overall tolerability was good. No vaccine-related serious adverse events occurred. Across the 24-month period, 53 SAEs were reported and were evenly distributed between arms. These findings are consistent with our earlier safety data and with other trials of reduced-dose mRNA boosters.^1,8–11,16^ The COVE trial likewise reported no new safety signals after mRNA-1273 boosting, reinforcing the favourable safety profile of mRNA platforms over 24 months of follow-up.^29^

This study has several limitations. Suspected SARS-CoV-2 infections were inferred from increases in anti-spike IgG titres, using a ≥ 1·2 fold threshold without anti-nucleocapsid testing. This arbitrary threshold may have led to misclassification of SARS-CoV-2 exposure or reinfection.^15^ Reliance on self-reported testing likely underestimated symptomatic infections.^35^ We did not prespecify variance comparisons between priming cohorts; the wider dispersion seen in BBIBP-CorV–primed sVNT values should therefore be interpreted cautiously. Neutralising activity was assessed only against the ancestral strain and Omicron BA.1. The immunological assays used have inherent constraints. QuantiFERON IGRA measures IFN-γ release from stimulated whole blood but does not define T-cell subsets, polyfunctionality, or mucosal responses, and ELISA- and sVNT assays similarly provide limited resolution of immune breadth.^36^ While analyses of binding antibody responses to JN.1 and other currently circulating Omicron sublineages are underway, they are not included here. Findings may not be generalisable to older adults, immunocompromised individuals, or populations with different variant exposure histories or vaccine access. However, retention remained high and serological data were available for most participants, supporting the robustness of long-term immunogenicity estimates.

## Conclusion

This trial provides comprehensive 24-month immunogenicity and safety data from a low-resource setting. With 87% retention, detailed serological follow-up, and head-to-head comparisons across three non-mRNA priming regimens, no COVID-19–related hospitalisations or deaths occurred despite evidence of undocumented exposure. Neutralising activity was sustained and IFN-γ responses remained detectable, while binding IgG waned towards baseline. Programmatically, fractional (15 µg) BNT162b2 performed comparably to standard (30 µg) dosing with reassuring safety, supporting fractional dosing as an evidence-based, cost-saving option to extend supply, widen coverage, and improve equity in booster campaigns, particularly in resource-constrained settings, while scheduling remains responsive to local epidemiology and variant antigenicity.

## Contributors

TBa, KMu, CvM, HT, BO, PVL, US, KBa, and GD conceptualised the study. KMu, TBa, and HT acquired funding. TBa, HT, BT, and FJ managed and coordinated the project. TBa, HT, OA, KJ, LB, and OJ recruited the participants, did the investigations, and collected the data. PVL, NM, KMa, SL, LAHD, BA, SJ, TBu, NA and NN managed sample collection and laboratory analyses. HT, FJ, and JDH monitored the trial. EFGN managed and curated the databases.

KAM and CDN wrote the statistical analysis plan. EFGN analysed the data. EFGN wrote the original draft of the manuscript. HT, CvM, FJ, and KBr provided supervision and oversight. JDH was the medical adviser. All authors reviewed the manuscript. All authors had full access to all the data in the study and had final responsibility for the decision to submit for publication.

## Data sharing statement

Anonymous participant data that underlie the results reported in this Article will be available on completion of the clinical trial. Data requests should be sent to the corresponding author. The requester must provide a scientifically sound proposal and data transfer agreement for the sponsors’ and collaborators’ approval. On approval, data will be transferred through a secure online platform.

## Data Availability

All data produced in the present study are available upon reasonable request to the authors.

https://pubmed.ncbi.nlm.nih.gov/38357398/

https://www.medrxiv.org/content/10.1101/2025.08.29.25334703v1

## Acknowledgements

This study was made possible by the generosity of the study participants – we thank them for their time and cooperation in the study procedures. We would like to acknowledge the invaluable contributions of: Dr Chinburen Jigjidsuren (Member of Parliament); Dr Ganzorig Dorjdagva (Ministry of Health); Dr Urangoo Khurlee, Dr Gentsenpilmaa Batbold, Dr Gantuya Damdinsuren, and Dr Altanshahai Boldbaatar (First Central Hospital of Mongolia); Dr Tserendagva Dalkh (Mongolian National University of Medical Sciences); Dr Davaalkham Jagdagsuren, Dr Naranzul Tsedenbal, Tserendulam Bazarkhuu, and Ariunbileg Gankhuyag (National Centre for Communicable Diseases); Dr Budkhand Ichinkhorloo, and Dr Erdenebayar Tsedevsuren (Onoshmed Laboratory); Dr Baldandugar Zeeren and Dr Davaajav Tsend-Ayush (Bayangol District Health Department); Bat-Ireedui Purevbaatar and Dr Gantuya Gansukh (Sukhbaatar District Health Department); Dr Davaajargal Oyunsuren (Songinokhairkhan District Health Department); Dr Gandiimaa Riimaadai, Dr Tserendulam Dorjsuren and Dr Ariunaa Jadambaa (Arkhangai Province Health Department); and Dr Erkegul Sandalhan (Railway Central Hospital). This clinical trial is funded by CEPI, an innovative global partnership working to accelerate the development of vaccines against epidemic and pandemic threats so they can be accessible to all people in need. Grant management support is provided by PATH, an international non-profit global health organization. The Government of Mongolia provided the study vaccines. Kerryn Moore is supported by an Australian National Health and Medical Research Council (NHMRC) Early Career Fellowship (Grant Number APP1160936); the content of this publication is solely the responsibility of the authors and does not necessarily represent the official views of the NHMRC. Authors affiliated with MCRI were supported by the Victorian Government’s Operational Infrastructure Support Program.

## References

1. Nanthapisal S, Puthanakit T, Jaru-Ampornpan P, et al. A randomized clinical trial of a booster dose with low versus standard dose of AZD1222 in adult after 2 doses of inactivated vaccines. Vaccine 2022; 40(18): 2551–60.

2. Niyomnaitham S, Jongkaewwattana A, Meesing A, et al. Immunogenicity of a fractional or full third dose of AZD1222 vaccine or BNT162b2 messenger RNA vaccine after two doses of CoronaVac vaccines against the Delta and Omicron variants. Int J Infect Dis 2023; 129: 19–31.

3. Liu X, Munro APS, Feng S, et al. Persistence of immunogenicity after seven COVID-19 vaccines given as third dose boosters following two doses of ChAdOx1 nCov-19 or BNT162b2 in the UK: Three month analyses of the COV-BOOST trial. J Infect 2022; 84(6): 795–813.

4. Liu X, Munro APS, Wright A, et al. Persistence of immune responses after heterologous and homologous third COVID-19 vaccine dose schedules in the UK: eight-month analyses of the COV-BOOST trial. J Infect 2023; 87(1): 18–26.

5. Steenackers K, Hanning N, Bruckers L, et al. Humoral immune response against SARS-CoV-2 after adapted COVID-19 vaccine schedules in healthy adults: The IMCOVAS randomized clinical trial. Vaccine 2024; 42(25): 126117.

6. Barros Verruck J, Moreira Puga MA, de Oliveira RD, et al. Antispike IgG antibody decay after immunisation with fractional versus full booster doses of COVID-19 vaccines: a 6-month longitudinal analysis of the FRACT-COV trial in Brazil. BMJ Public Health 2025; 3(2): e002331.

7. Wiecek W, Ahuja A, Chaudhuri E, et al. Testing fractional doses of COVID-19 vaccines. Proc Natl Acad Sci U S A 2022; 119(8).

8. Batmunkh T, Moore KA, Thomson H, et al. Immunogenicity, safety, and reactogenicity of a half- versus full-dose BNT162b2 (Pfizer-BioNTech) booster following a two-dose ChAdOx1 nCoV-19, BBIBP-CorV, or Gam-COVID-Vac priming schedule in Mongolia: a randomised, controlled, non-inferiority trial. Lancet Reg Health West Pac 2024; 42: 100953.

9. Batmunkh T, Neal EFG, Amraa O, et al. Twelve-month follow-up of immunogenicity and safety of fractional and standard booster doses of the Pfizer-BioNTech COVID-19 vaccine in adults primed with ChAdOx1, BBIBP-CorV, or GAM-CoV-Vac: a randomised controlled trial. medRxiv 2025: 2025.08.29.25334703.

10. Munro APS, Janani L, Cornelius V, et al. Safety and immunogenicity of seven COVID-19 vaccines as a third dose (booster) following two doses of ChAdOx1 nCov-19 or BNT162b2 in the UK (COV-BOOST): a blinded, multicentre, randomised, controlled, phase 2 trial. Lancet 2021; 398(10318): 2258–76.

11. Fadlyana E, Setiabudi D, Kartasasmita CB, et al. Immunogenicity and safety in healthy adults of full dose versus half doses of COVID-19 vaccine (ChAdOx1-S or BNT162b2) or full-dose CoronaVac administered as a booster dose after priming with CoronaVac: a randomised, observer-masked, controlled trial in Indonesia. Lancet Infect Dis 2023; 23(5): 545–55.

12. Moreira Puga MA, Dias de Oliveira R, Vieira da Silva P, et al. Immunogenicity and reactogenicity of fractional vs. full booster doses of COVID-19 vaccines: a non-inferiority, randomised, double-blind, phase IV clinical trial in Brazil. Lancet Reg Health Am 2025; 44: 101031.

13. Arunachalam PS, Lai L, Samaha H, et al. Durability of immune responses to mRNA booster vaccination against COVID-19. J Clin Invest 2023; 133(10).

14. Khoury DS, Docken SS, Subbarao K, Kent SJ, Davenport MP, Cromer D. Predicting the efficacy of variant-modified COVID-19 vaccine boosters. Nat Med 2023; 29(3): 574–8.

15. Branche AR, Rouphael NG, Losada C, et al. Immunogenicity of the BA.1 and BA.4/BA.5 Severe Acute Respiratory Syndrome Coronavirus 2 Bivalent Boosts: Preliminary Results From the COVAIL Randomized Clinical Trial. Clin Infect Dis 2023; 77(4): 560–4.

16. Cheng H, Peng Z, Si S, et al. Immunogenicity and Safety of Homologous and Heterologous Prime-Boost Immunization with COVID-19 Vaccine: Systematic Review and Meta-Analysis. Vaccines (Basel) 2022; 10(5).

17. Sun P, Balinsky CA, Jiang L, et al. Antibody Responses to the SARS-CoV-2 Ancestral Strain and Omicron Variants in Moderna mRNA-1273 Vaccinated Active-Duty US Navy Sailors and Marines. J Infect Dis 2023; 228(2): 149–59.

18. Burbelo PD, Riedo FX, Morishima C, et al. Sensitivity in Detection of Antibodies to Nucleocapsid and Spike Proteins of Severe Acute Respiratory Syndrome Coronavirus 2 in Patients With Coronavirus Disease 2019. J Infect Dis 2020; 222(2): 206–13.

19. Seki Y, Yoshihara Y, Nojima K, et al. Safety and immunogenicity of the Pfizer/BioNTech SARS-CoV-2 mRNA third booster vaccine dose against the BA.1 and BA.2 Omicron variants. Med 2022; 3(6): 406–21 e4.

20. Fujitani M, Lu X, Shinnakasu R, et al. Longitudinal analysis of immune responses to SARS-CoV-2 recombinant vaccine S-268019-b in phase 1/2 prime-boost study. Front Immunol 2025; 16: 1550279.

21. Harris PA, Taylor R, Minor BL, et al. The REDCap consortium: Building an international community of software platform partners. J Biomed Inform 2019; 95: 103208.

22. Harris PA, Taylor R, Thielke R, Payne J, Gonzalez N, Conde JG. Research electronic data capture (REDCap)--a metadata-driven methodology and workflow process for providing translational research informatics support. J Biomed Inform 2009; 42(2): 377–81.

23. International Council for Harmonisation of Technical Requirements for Pharmaceuticals for Human Use. Addendum on estimands and sensitivity analysis in clinical trials to the guideline on statistical principles for clinical trials. E9(R1); 2019. p. 20.

24. Rubin DB. Multiple imputation for survey nonresponse. New York: Wiley; 1987.

25. White IR, Royston P, Wood AM. Multiple imputation using chained equations: Issues and guidance for practice. Stat Med 2011; 30(4): 377–99.

26. Hart JD, Fadlyana E, Mazarakis N, et al. Immunogenicity of fractional and standard dose COVID-19 vaccine boosters among healthy adults in Indonesia: twenty four month follow-up from a randomised controlled trial. Nat Commun 2025; In Press.

27. StataCorp. Stata Statistical Software. Release 18.5. ed. College Station, TX: StataCorp LLC; 2024.

28. Batmunkh T, Neal EFG, Amraa O, et al. Twelve-month follow-up of immunogenicity and safety of fractional and standard booster doses of the Pfizer-BioNTech COVID-19 vaccine in adults primed with ChAdOx1, BBIBP-CorV, or GAM-CoV-Vac: a randomised controlled trial. Submitted for publication - under review 2025.

29. Baden LR, El Sahly HM, Essink B, et al. Long-term safety and effectiveness of mRNA-1273 vaccine in adults: COVE trial open-label and booster phases. Nat Commun 2024; 15(1): 7469.

30. Korosec CS, Farhang-Sardroodi S, Dick DW, et al. Long-term durability of immune responses to the BNT162b2 and mRNA-1273 vaccines based on dosage, age and sex. Sci Rep 2022; 12(1): 21232.

31. Prakash S, Dhanushkodi NR, Zayou L, et al. Cross-Protection Induced by Highly Conserved Human B, CD4(+,) and CD8(+) T Cell Epitopes-Based Coronavirus Vaccine Against Severe Infection, Disease, and Death Caused by Multiple SARS-CoV-2 Variants of Concern. bioRxiv 2023.

32. Tarke A, Ramezani-Rad P, Alves Pereira Neto T, et al. SARS-CoV-2 breakthrough infections enhance T cell response magnitude, breadth, and epitope repertoire. Cell Rep Med 2024; 5(6): 101583.

33. Sankaranarayanan S, Mohkhedkar M, Janakiraman V. Mutations in spike protein T cell epitopes of SARS-COV-2 variants: Plausible influence on vaccine efficacy. Biochim Biophys Acta Mol Basis Dis 2022; 1868(9): 166432.

34. Tschiderer L, Innerhofer H, Seekircher L, et al. Long-term effectiveness of an ultra-rapid rollout vaccination campaign with BNT162b2 on the incidence of SARS-CoV-2 infection. iScience 2024; 27(11): 111117.

35. Anand A, Vialard F, Esmail A, et al. Self-tests for COVID-19: What is the evidence? A living systematic review and meta-analysis (2020-2023). PLOS Glob Public Health 2024; 4(2): e0002336.

36. Johnson SA, Phillips E, Adele S, et al. Evaluation of QuantiFERON SARS-CoV-2 interferon-γ release assay following SARS-CoV-2 infection and vaccination. Clin Exp Immunol 2023; 212(3): 249–61.

