## Supplementary Materials for "Fractional versus standard BNT162b2 boosters after non-mRNA priming in Mongolia: 24-month immunogenicity and safety evidence from a randomised controlled trial"

### Immunological responses up to 24 months post-booster

#### *Missingness of anti-spike IgG and sVNT inhibition*

Missing data for anti-spike IgG and surrogate virus neutralisation test (sVNT) inhibition are summarised in Supplementary Table 1. IgG missingness was low at early timepoints: 0·3% (2/601) at baseline, 2·3% (14/601) at day 28, 4·5% (27/601) at six months, and 4·8% (29/601) at 12 months. Missingness increased at later visits, with 12·0% (72/601) at 18 months and 13·5% (81/601) at 24 months. For sVNT, missingness followed the same pattern, and proportions were identical between Wuhan-Hu-1 and Omicron (BA.1) at every visit (baseline 0·3%, day 28 2·3%, six months 4·5%, 12 months 5·6%, 18 months 12·0%, 24 months 13·5%). All missing data at 24 months were due to participant withdrawal or loss to follow-up, with similar rates across study arms and priming strata.

When stratified by study arm, missingness at each time point was similar between the standard and fractional dose groups. At 24 months, IgG was missing for 13·3% (40/300) in the standard group and 13·6% (41/301) in the fractional group, indicating no systematic differences in visit attendance or data availability by dosing strategy. Patterns were also similar across priming vaccine strata; at 24 months, missingness ranged from 10·8% to 15·0% across ChAdOx1-S, BBIBP-CorV and Gam-COVID-Vac groups, with no difference in the missingness rate across priming strata.

Covariate data used in regression models were nearly complete. Age group, priming vaccine, and dates of doses 1 and 2 had 0·0% missingness; the date of the third (study) dose was missing for 3/601 (0·5%) participants who did not receive their allocated study vaccine.

*Supplementary Table 1 Missingness of anti-spike IgG and sVNT inhibition (Wuhan-Hu-1 and Omicron BA.1) at baseline, day 28, six, 12, 18, and 24 months, overall and stratified by study arm and priming vaccine.*

| | All Priming Strata | | | Primed with ChAdOx1-S | | Primed with BBIBP-CorV | | Primed with Gam-COVID-Vac | |
|---|---|---|---|---|---|---|---|---|---|
| Variable | Total (N = 601) | Standard (N = 300) | Fractional (N = 301) | Standard (N = 65) | Fractional (N = 65) | Standard (N = 201) | Fractional (N = 200) | Standard (N = 34) | Fractional (N = 36) |
| | n (%) | n (%) | n (%) | n (%) | n (%) | n (%) | n (%) | n (%) | n (%) |
| Anti-spike IgG levels | | | | | | | | | |
| Baseline | 2 (0·3) | 1 (0·3) | 1 (0·3) | 0 (0·0) | 1 (1·5) | 1 (0·5) | 0 (0·0) | 0 (0·0) | 0 (0·0) |
| Day 28 | 14 (2·3) | 8 (2·7) | 6 (2·0) | 0 (0·0) | 3 (4·6) | 7 (3·5) | 2 (1·0) | 1 (2·9) | 1 (2·8) |
| Six months | 27 (4·5) | 16 (5·3) | 11 (3·7) | 5 (7·7) | 5 (7·7) | 9 (4·5) | 5 (2·5) | 2 (5·9) | 1 (2·8) |
| 12 months | 29 (4·8) | 15 (5·0) | 14 (4·7) | 4 (6·2) | 4 (6·2) | 9 (4·5) | 9 (4·5) | 2 (5·9) | 1 (2·8) |
| 18 months | 72 (12·0) | 36 (12·0) | 36 (12·0) | 9 (13·8) | 7 (10·8) | 23 (11·4) | 27 (13·5) | 4 (11·8) | 2 (5·6) |
| 24 months | 81 (13·5) | 40 (13·3) | 41 (13·6) | 7 (10·8) | 7 (10·8) | 29 (14·4) | 30 (15·0) | 4 (11·8) | 4 (11·1) |
| sVNT inhibition (Wuhan-Hu-1 and Omicron BA.1) | | | | | | | | | |
| Baseline | 2 (0·3) | 1 (0·3) | 1 (0·3) | 0 (0·0) | 1 (1·5) | 1 (0·5) | 0 (0·0) | 0 (0·0) | 0 (0·0) |
| Day 28 | 14 (2·3) | 8 (2·7) | 6 (2·0) | 0 (0·0) | 3 (4·6) | 7 (3·5) | 2 (1·0) | 1 (2·9) | 1 (2·8) |
| Six months | 27 (4·5) | 16 (5·3) | 11 (3·7) | 5 (7·7) | 5 (7·7) | 9 (4·5) | 5 (2·5) | 2 (5·9) | 1 (2·8) |

| | | | | | | | | | |
|---|---|---|---|---|---|---|---|---|---|
| 12 months | 34 (5·7) | 18 (6·0) | 16 (5·3) | 4 (6·2) | 4 (6·2) | 11 (5·5) | 10 (5·0) | 3 (8·8) | 2 (5·6) |
| 18 months | 72 (12·0) | 36 (12·0) | 36 (12·0) | 9 (13·8) | 7 (10·8) | 23 (11·4) | 27 (13·5) | 4 (11·8) | 2 (5·6) |
| 24 months | 81 (13·5) | 40 (13·3) | 41 (13·6) | 7 (10·8) | 7 (10·8) | 29 (14·4) | 30 (15·0) | 4 (11·8) | 4 (11·1) |
| Baseline covariates[a] | | | | | | | | | |
| Age group | 0 (0·0) | 0 (0·0) | 0 (0·0) | 0 (0·0) | 0 (0·0) | 0 (0·0) | 0 (0·0) | 0 (0·0) | 0 (0·0) |
| Priming vaccine | 0 (0·0) | 0 (0·0) | 0 (0·0) | 0 (0·0) | 0 (0·0) | 0 (0·0) | 0 (0·0) | 0 (0·0) | 0 (0·0) |
| Date dose 1 received | 0 (0·0) | 0 (0·0) | 0 (0·0) | 0 (0·0) | 0 (0·0) | 0 (0·0) | 0 (0·0) | 0 (0·0) | 0 (0·0) |
| Date dose 2 received | 0 (0·0) | 0 (0·0) | 0 (0·0) | 0 (0·0) | 0 (0·0) | 0 (0·0) | 0 (0·0) | 0 (0·0) | 0 (0·0) |
| Date dose 3 received[b] | 3 (0·5) | 1 (0·3) | 2 (0·3) | 0 (0·0) | 1 (1·5) | 1 (0·5) | 1 (0·5) | 0 (0·0) | 0 (0·0) |
| Study day of blood draw | | | | | | | | | |
| Baseline | 1 (0·2) | 1 (0·3) | 0 (0·0) | 0 (0·0) | 0 (0·0) | 1 (0·5) | 0 (0·0) | 0 (0·0) | 0 (0·0) |
| Day 28 | 14 (2·3) | 8 (2·7) | 6 (2·0) | 0 (0·0) | 3 (4·6) | 7 (3·5) | 2 (1·0) | 1 (2·9) | 1 (2·8) |
| Six months | 27 (4·5) | 16 (5·3) | 11 (3·7) | 5 (7·7) | 5 (7·7) | 9 (4·5) | 5 (2·5) | 2 (5·9) | 1 (2·8) |
| 12 months | 29 (4·8) | 15 (5·0) | 14 (4·7) | 4 (6·2) | 4 (6·2) | 9 (4·5) | 9 (4·5) | 2 (5·9) | 1 (2·8) |
| 18 months | 72 (12·0) | 36 (12·0) | 36 (12·0) | 9 (13·8) | 7 (10·8) | 23 (11·4) | 27 (13·5) | 4 (11·8) | 2 (5·6) |
| 24 months | 81 (13·5) | 40 (13·3) | 41 (13·6) | 7 (10·8) | 7 (10·8) | 29 (14·4) | 30 (15·0) | 4 (11·8) | 4 (11·1) |

**Abbreviations:** BA.1 - Omicron BA.1 subvariant; IgG - immunoglobulin G; N - total number of participants; n - number with missing data; sVNT - surrogate virus neutralisation test

**Footnotes**: [a] Covariate completeness at baseline is shown for age group, priming vaccine, and dates of doses 1–3; [b] Date of dose 3 is missing only for participants who did not receive their allocated study booster

*Missingness of 24 months anti-spike IgG and sVNT inhibition by baseline characteristics*

To explore whether missingness of anti-spike IgG and sVNT inhibition at 24 months was systematically associated with participant characteristics, we summarised 24 month missing immune-response data by baseline covariates, stratified by study arm and priming vaccine (Supplementary Table 2). Overall, missingness was higher among participants aged 18–<50 years (17·5%) than those ≥50 years (7·5%), and among males (15·7%) compared with females (11·6%). Differences across comorbidity groups were small in absolute terms and showed no consistent pattern. Rates were broadly similar between the standard-dose and fractional-dose groups and across priming strata (ChAdOx1-S, BBIBP-CorV, Gam-COVID-Vac), providing no evidence of differential attrition by treatment allocation or baseline health status.

*Supplementary Table 2 Missingness of anti-spike IgG and sVNT inhibition at 24 months, stratified by baseline characteristics, study arm, and priming vaccine[a]*

| Variable | All Priming Strata | | | Primed with ChAdOx1-S | | Primed with BBIBP-CorV | | Primed with Gam-COVID-Vac | |
|---|---|---|---|---|---|---|---|---|---|
| | Total (N = 601) | Standard (N = 300) | Fractional (N = 300) | Standard (N = 65) | Fractional (N = 65) | Standard (N = 201) | Fractional (N = 200) | Standard (N = 34) | Fractional (N = 36) |
| 18–<50 years of age | 63/361 (17·5) | 33/181 (18·2) | 30/180 (16·7) | 7/54 (13·0) | 5/51 (9·8) | 24/103 (23·3) | 22/103 (21·4) | 2/24 (8·3) | 3/26 (11·5) |
| ≥50 years of age | 18/240 (7·5) | 7/119 (5·9) | 11/121 (9·1) | 0/11 (0·0) | 2/14 (14·3) | 5/98 (5·1) | 8/97 (8·3) | 2/10 (20·0) | 1/10 (10·0) |
| Male sex | 43/274 (15·7) | 17/132 (12·8) | 26/141 (18·4) | 5/32 (15·6) | 3/33 (9·1) | 10/86 (11·6) | 19/86 (22·1) | 2/15 (13·3) | 4/22 (18·2) |
| Female sex | 38/327 (11·6) | 23/167 (13·8) | 15/160 (9·4) | 2/33 (6·1) | 4/32 (12·5) | 19/115 (16·5) | 11/114 (9·7) | 2/19 (10·5) | 0/14 (0·0) |
| BMI, kg/m$^2$, median (IQR) | 23·9 (21·7-26) | 24·1 (21·8-5·7) | 23·6 (21·6-26·7) | 26·2 (23·9-31·1) | 23·1 (21·5-24·8) | 22·9 (21·5-24·5) | 23·5 (21·5-26·8) | 25·1 (24·2-26·3) | 27·4 (25·2-32·1) |
| Days between 1st and 2nd doses, median (IQR) | 30 (24-44) | 32 (25·5-49) | 30 (23-43) | 43 (41-53) | 44 (38-49) | 28 (24-39) | 29 (23-37) | 53·5 (35-68·5) | 60 (38·5-72) |
| Days between 2nd and study (3rd) dose, median (IQR) | 409 (383-454) | 389·5 (368·5-452) | 425 (403-463) | 505 (493-528) | 494 (404-526) | 377 (365-392) | 417 (400-446) | 445 (425-474) | 339 (407·5-455·5) |
| Reaction following 1st or 2nd dose | 8/93 (8·6) | 5/45 (11·1) | 3/48 (6·3) | 2/21 (9·5) | 0/18 (0·0) | 3/17 (17·7) | 2/24 (8·3) | 0/7 (0·0) | 1/6 (16·7) |
| Pain or fever medication for reaction | 0/21 (0·0) | 0/9 (0·0) | 0/12 (0·0) | 0/7 (0·0) | 0/8 (0·0) | 0/1 (0·0) | 0/4 (0·0) | 0/1 (0·0) | NA |
| Medical advice sought for reaction | 0/3 (0·0) | 0/1 (0·0) | 0/2 (0·0) | 0/1 (0·0) | 0/1 (0·0) | NA | 0/1 (0·0) | NA | NA |
| Symptoms of reaction resolved | 5/53 (9·4) | 4/28 (14·3) | 1/25 (4·0) | 2/11 (18·2) | 0/5 (0·0) | 2/10 (20·0) | 0/15 (0·0) | 0/7 (0·0) | 1/5 (20·0) |
| Self-reported prior SARS-CoV-2 | 31/294 (10·5) | 14/150 (9·3) | 17/144 (11·8) | 7/39 (18·0) | 2/41 (4·9) | 6/91 (6·6) | 11/74 (14·9) | 1/20 (5·0) | 4/29 (13·8) |
| Comorbidities | | | | | | | | | |
| Obesity (BMI ≥30 kg/m$^2$) | 9/115 (7·8) | 3/55 (5·5) | 6/60 (10·0) | 2/13 (15·4) | 0/12 (0·0) | 1/37 (2·7) | 5/39 (12·8) | 0/5 (0·0) | 1/9 (11·1) |
| Diabetes mellitus | 2/25 (8·0) | 2/17 (11·8) | 0/8 (0·0) | 0/2 (0·0) | 0/1 (0·0) | 1/10 (10·0) | 0/5 (0·0) | 1/5 (20·0) | 0/2 (0·0) |
| Cardiovascular disease | 5/56 (8·9) | 1/26 (3·9) | 4/30 (13·3) | 0/2 (0·0) | 0/5 (0·0) | 0/20 (0·0) | 4/23 (17·4) | 1/4 (25·0) | 0/2 (0·0) |
| Hypertension | 16/168 (9·5) | 6/81 (7·4) | 10/87 (11·5) | 2/14 (14·3) | 2/15 (13·3) | 3/60 (5·0) | 6/60 (10·0) | 1/7 (14·3) | 2/12 (16·7) |
| Cancer | 1/3 (33·3) | NA | 1/3 (33·3) | NA | NA | NA | 1/1 (100·0) | NA | 0/2 (0·0) |
| COPD | 0/8 (0·0) | 0/4 (0·0) | 0/4 (0·0) | NA | NA | 0/2 (0·0) | 0/3 (0·0) | 0/2 (0·0) | 0/1 (0·0) |
| Chronic kidney disease | 4/49 (8·2) | 1/25 (4·0) | 3/24 (12·5) | 0/3 (0·0) | 0/4 (0·0) | 1/18 (5·6) | 3/17 (17·6) | 0/4 (0·0) | 0/3 (0·0) |
| Chronic liver disease | 1/19 (5·3) | 1/9 (11·1) | 0/10 (0·0) | 0/1 (0·0) | 0/3 (0·0) | 1/6 (16·7) | 0/4 (0·0) | 0/2 (0·0) | 0/3 (0·0) |
| History of anaphylaxis | 0/12 (0·0) | 0/6 (0·0) | 0/6 (0·0) | 0/3 (0·0) | 0/2 (0·0) | 0/3 (0·0) | 0/3 (0·0) | NA | 0/1 (0·0) |
| Neurological disease | 1/6 (16·7) | 0/3 (0·0) | 1/3 (33·3) | 0/1 (0·0) | 0/1 (0·0) | 0/2 (0·0) | 1/2 (50·0) | NA | NA |
| On anticoagulant therapy | 2/33 (6·1) | 1/17 (5·9) | 1/16 (6·3) | 0/4 (0·0) | 0/2 (0·0) | 0/9 (0·0) | 1/11 (9·1) | 1/4 (25·0) | 0/3 (0·0) |
| Immunocompromised | NA | NA | NA | NA | NA | NA | NA | NA | NA |
| Mastocytosis causing recurrent anaphylaxis | 1/1 (100·0) | NA | 1/1 (100·0) | NA | NA | NA | 1/1 (100·0) | NA | NA |
| Cigarette user | 15/125 (12·0) | 8/66 (22·1) | 7/59 (11·9) | 1/16 (6·3) | 1/17 (5·9) | 4/37 (10·8) | 6/34 (17·7) | 3/13 (23·1) | 0/8 (0·0) |
| Currently pregnant | 0/1 (0·0) | 0/1 (0·0) | NA | 0/1 (0·0) | NA | NA | NA | NA | NA |

**Abbreviations:** BMI - body mass index; IQR - interquartile range; N - total number of participants; n - number with missing immunogenicity data; NA - not applicable (no participants in that stratum).

**Footnotes:** [a] Data are presented as n/N (%), unless otherwise indicated.

#### *Anti-spike IgG levels*

Supplementary Table 3 summarises the geometric mean concentration (GMC) with 95% confidence interval (CI) of anti-spike IgG antibodies in each study arm and the geometric mean ratio (GMR) with 95% CI comparing the fractional and standard dose arms at all time points.

*Supplementary Table 3 Geometric mean concentration of anti-spike IgG and geometric mean ratio (fractional vs standard dose) at baseline, day 28, six, 12, 18, and 24 months, overall and stratified by priming vaccine*

| Priming Strata | GMC IgG (95% CI) | | GMR fractional/standard (95% CI) |
|---|---|---|---|
| | Standard Dose | Fractional Dose | |
| Baseline[a] | | | |
| All | 969 (876, 1072) [n = 299] | 929 (839, 1029) [n = 300] | 0·94 (0·82, 1·08); p = 0·398 |
| ChAdOx1-S | 1023 (845, 1238) [n = 65] | 958 (820, 1119) [n = 64] | 0·94 (0·74, 1·20); p = 0·620 |
| BBIBP-CorV | 956 (843, 1084) [n = 200] | 899 (785, 1031) [n = 200] | 0·92 (0·77, 1·10); p = 0·370 |
| Gam-COVID-Vac | 953 (667, 1360) [n = 34] | 1052 (788, 1404) [n = 36] | 1·09 (0·72, 1·65); p = 0·685 |
| Day 28[b] | | | |
| All | 4946 (4614, 5302) [n = 292] | 4619 (4292, 4970) [n = 295] | 0·95 (0·87, 1·05); p = 0·307 |
| ChAdOx1-S | 4394 (3863, 4997) [n = 65] | 4167 (3550, 4892) [n = 62] | 0·96 (0·79, 1·16); p = 0·652 |
| BBIBP-CorV | 5109 (4678, 5580) [n = 194] | 4970 (4541, 5438) [n = 198] | 0·99 (0·88, 1·12); p = 0·891 |
| Gam-COVID-Vac | 5160 (4128, 6450) [n = 33] | 3662 (3016, 4447) [n = 35] | 0·72 (0·55, 0·94); p = 0·016 |
| Six months[b] | | | |
| All | 2085 (1926, 2256) [n = 284] | 2123 (1964, 2296) [n = 290] | 1·03 (0·93, 1·15); p = 0·586 |
| ChAdOx1-S | 1832 (1556, 2157) [n = 60] | 1706 (1466, 1985) [n = 60] | 0·93 (0·76, 1·15); p = 0·519 |
| BBIBP-CorV | 2195 (1991, 2419) [n = 192] | 2286 (2074, 2521) [n = 195] | 1·05 (0·92, 1·20); p = 0·492 |
| Gam-COVID-Vac | 1950 (1511, 2517) [n = 32] | 2044 (1628, 2569) [n = 35] | 1·09 (0·79, 1·51); p = 0·588 |
| 12 months[b] | | | |
| All | 2204 (2030, 2393) [n = 285] | 2208 (2027, 2404) [n = 287] | 1·01 (0·90, 1·13); p = 0·891 |
| ChAdOx1-S | 2270 (1952, 2639) [n = 61] | 1765 (1524, 2045) [n = 61] | 0·79 (0·65, 0·98); p = 0·028 |
| BBIBP-CorV | 2208 (1990, 2449) [n = 192] | 2310 (2072, 2574) [n = 191] | 1·04 (0·90, 1·21); p = 0·557 |
| Gam-COVID-Vac | 2063 (1553, 2740) [n = 32] | 2547 (1957, 3313) [n = 35] | 1·19 (0·83, 1·71); p = 0·331 |
| 18 months[b] | | | |
| All | 1336 (1231, 1450) [n = 264] | 1425 (1313, 1546) [n = 265] | 1·08 (0·97, 1·22); p = 0·156 |
| ChAdOx1-S | 1344 (1129, 1599) [n = 56] | 1192 (1027, 1383) [n = 58] | 0·90 (0·72, 1·12); p = 0·352 |
| BBIBP-CorV | 1365 (1231, 1514) [n = 178] | 1498 (1348, 1666) [n = 173] | 1·11 (0·96, 1·28); p = 0·161 |
| Gam-COVID-Vac | 1166 (932, 1459) [n = 30] | 1495 (1186, 1886) [n = 34] | 1·30 (0·96, 1·77); p = 0·089 |
| 24 months[b] | | | |
| All | 1051 (973, 1135) [n = 260] | 1107 (1021, 1199) [n = 260] | 1·06 (0·95, 1·18); p = 0·271 |
| ChAdOx1-S | 931 (807, 1075) [n = 58] | 1031 (901, 1179) [n = 58] | 1·13 (0·94, 1·37); p = 0·183 |
| BBIBP-CorV | 1101 (995, 1217) [n = 172] | 1088 (982, 1206) [n = 170] | 1·00 (0·87, 1·15); p = 0·954 |
| Gam-COVID-Vac | 1018 (835, 1240) [n = 30] | 1376 (1048, 1806) [n = 32] | 1·28 (0·91, 1·80); p = 0·153 |

**Abbreviations:** CI - confidence interval; GMR - geometric mean ratio; IgG - immunoglobulin G. **Footnotes:** [a] At baseline, the GMR is adjusted for age group, priming vaccine, interval between doses 1–2, interval between dose 2 and the study dose, and study day of blood draw; [b] At day 28, six, 12, 18, and 24 months, the GMR is adjusted for the same variables and baseline anti-spike IgG.

#### *Multiple imputation*

As prespecified in the statistical analysis plan, multiple imputation by chained equations (MICE) was undertaken for the primary immunogenicity outcome (anti-spike IgG). Data were reshaped to wide format with fixed time points (day 28, six, 12, 18 and 24 months). Imputations were run separately by study arm to preserve treatment effects, using chained linear regression with 50 imputations (seed 15072025). The imputation model included age group, priming vaccine, study day of blood draw, days between doses 1–2, days between dose 2 and the study dose, and baseline log-IgG. GMCs were obtained by exponentiating MI-pooled means, and GMRs were estimated from MI-pooled linear regression models on the log scale, adjusted for baseline log-IgG and covariates. Estimates and standard errors were combined across imputations using Rubin's rules.[1,2] Supplementary Table 4 summarises the GMC (95% CI) of anti-spike IgG antibodies in each study arm and the GMR (95% CI) comparing the fractional and standard dose arms at all time points, using MICE to account for missing data.

*Supplementary Table 4 Geometric mean concentration of anti-spike IgG and geometric mean ratio (fractional vs standard dose) at baseline, day 28, six, 12, 18, and 24 months, overall and stratified by priming vaccine, with missing data imputed using MICE[a]*

| Time point | GMC IgG (95% CI) | | GMR fractional/standard (95% CI) |
|---|---|---|---|
| | Standard Dose | Fractional Dose | |
| Day 28 | 4946 (4616, 5300) [n = 300] | 4620 (4296, 4969) [n = 301] | 0·95 (0·86 - 1·04); p = 0·287 |
| Six months | 2072 (1916, 2239) [n = 300] | 2118 (1959, 2290) [n = 301] | 1·03 (0·93 - 1·15); p = 0·540 |
| 12 months | 2203 (2030, 2391) [n = 300] | 2201 (2023, 2395) [n = 301] | 1·01 (0·90 - 1·13); p = 0·896 |
| 18 months | 1341 (1238, 1453) [n = 300] | 1411 (1301, 1529) [n = 301] | 1·07 (0·95 - 1·19); p = 0·255 |
| 24 months | 1043 (967, 1124) [n = 300] | 1074 (993, 1162) [n = 301] | 1·03 (0·93 - 1·15); p = 0·546 |

**Abbreviations:** CI - confidence interval; GMR - geometric mean ratio; IgG - immunoglobulin G. **Footnotes:** [a] Results are pooled across 50 imputed datasets generated using MICE; [b] At baseline, the GMR is adjusted for age group, priming vaccine, interval between doses 1–2, interval between dose 2 and the study dose, and study day of blood draw; [c] At day 28, six, 12, 18, and 24 months, the GMR is adjusted for the same variables and baseline anti-spike IgG.

Supplementary Table 5 presents anti-spike IgG GMCs (95% CIs) by study arm, stratified by age group (18–<50 years and ≥50 years) across all visits from baseline to 24 months.

*Supplementary Table 5 Geometric mean concentration of anti-spike IgG at baseline, day 28, six, 12, 18, and 24 months, stratified by age group and study arm*

| Priming Strata | GMC IgG (95% CI) | |
|---|---|---|
| | Standard Dose | Fractional Dose |
| Baseline | | |
| 18–<50 years | 798 (711, 896) [n = 181] | 799 (713, 896) [n = 179] |
| ≥50 years | 1307 (1099, 1554) [n = 118] | 1160 (966, 1393) [n = 121] |
| Day 28 | | |
| 18–<50 years | 4747 (4359, 5170) [n = 175] | 4313 (3930, 4733) [n = 175] |
| ≥50 years | 5259 (4673, 5918) [n = 117] | 5104 (4537, 3963) [n = 120] |
| Six months | | |
| 18–<50 years | 1890 (1711, 2087) [n = 170] | 1886 (1723, 2065) [n = 172] |
| ≥50 years | 2413 (2125, 2741) [n = 114] | 2523 (2205, 2887) [n = 118] |
| 12 months | | |
| 18–<50 years | 2124 (1922, 2348) [n = 173] | 2082 (1893, 2290) [n = 169] |
| ≥50 years | 2333 (2023, 2691) [n = 112] | 2400 (2052, 2807) [n = 118] |
| 18 months | | |
| 18–<50 years | 1336 (1231, 1451) [n = 153] | 1400 (1276, 1537) [n = 155] |
| ≥50 years | 1269 (1100, 1464) [n = 111] | 1460 (1258, 1694) [n = 110] |
| 24 months | | |
| 18–<50 years | 1049 (950, 1158) [n = 148] | 1034 (938, 1140) [n = 150] |
| ≥50 years | 1053 (930, 1193) [n = 112] | 1214 (1061, 1389) [n = 110] |

**Abbreviations:** CI - confidence interval; GMC - geometric mean concentration; IgG - immunoglobulin G.

#### *Missingness of IFN-γ Ag1 and Ag2*

Missingness of interferon-γ (IFN-γ) concentration data for Ag1 and Ag2 among participants enrolled in the cell-mediated immunity (CMI) subset is summarised in Supplementary Table 6. In the CMI subset, missingness for IFN-γ concentrations (Ag1 and Ag2; identical counts) was low at early timepoints (5·5% [14/256] at baseline, 7·0% [18/256] at day 28, 5·5% [14/256] at six months, and 5·9% [15/256] at 12 months), then increased at later visits (14·1% [36/256] at 18 months; 13·3% [34/256] at 24 months). Patterns were similar by study arm; at 24 months, missingness was 11·7% (15/128) in the standard-dose arm and 14·8% (19/128) in the fractional-dose arm. Across priming strata, 24-month missingness ranged from 5·3–17·2% (ChAdOx1-S: 12·0%/11·8% [standard/fractional]; BBIBP-CorV: 13·6%/17·2%; Gam-COVID-Vac: 5·3%/15·8%), noting small denominators for the Gam-COVID-Vac groups. Baseline covariates (age group, priming vaccine, and dates of COVID-19 vaccine doses one to three) were complete (0·0% missing). Documentation of the study day of blood draw showed higher missingness at later visits (13·7% [35/256] at 18 months; 13·3% [34/256] at 24 months overall), reflecting participant dropout at later visits.

*Supplementary Table 6 Missingness of IFN-γ concentrations for Ag1[a] and Ag2[b] at baseline, day 28, six, 12, 18, and 24 months in the cell-mediated immunity sub-study, overall and stratified by study arm and priming vaccine[c]*

| Variable | All Priming Strata | | | Primed with ChAdOx1-S | | Primed with BBIBP-CorV | | Primed with Gam-COVID-Vac | |
|---|---|---|---|---|---|---|---|---|---|
| | Total (N = 256) | Standard (N = 128) | Fractional (N= 128) | Standard (N = 50) | Fractional (N = 51) | Standard (N = 59) | Fractional (N = 58) | Standard (N = 19) | Fractional (N = 19) |
| | n/N (%) | n/N (%) | n/N (%) | n/N (%) | n/N (%) | n/N (%) | n/N (%) | n/N (%) | n/N (%) |
| IFN-γ concentrations | | | | | | | | | |
| Baseline | 14 (5·5) | 6 (4·7) | 8 (6·3) | 1 (2·0) | 3 (5·9) | 5 (8·5) | 5 (8·6) | 0 (0·0) | 0 (0·0) |
| Day 28 | 18 (7·0) | 8 (6·3) | 10 (7·8) | 1 (2·0) | 3 (5·9) | 6 (10·2) | 6 (10·3) | 1 (5·3) | 1 (5·3) |
| Six months | 14 (5·5) | 7 (5·5) | 7 (5·5) | 4 (8·0) | 4 (7·8) | 2 (3·4) | 2 (3·4) | 1 (5·3) | 1 (5·3) |
| 12 months | 15 (5·9) | 7 (5·5) | 8 (6·3) | 4 (8·0) | 3 (5·9) | 2 (3·4) | 4 (6·9) | 1 (5·3) | 1 (5·3) |
| 18 months | 36 (14·1) | 18 (14·1) | 18 (14·1) | 8 (16·0) | 6 (11·8) | 8 (13·6) | 11 (19·0) | 2 (10·5) | 1 (5·3) |
| 24 months | 34 (13·3) | 15 (11·7) | 19 (14·8) | 6 (12·0) | 6 (11·8) | 8 (13·6) | 10 (17·2) | 1 (5·3) | 3 (15·8) |
| Baseline covariates[c] | | | | | | | | | |
| Age group | 0 (0·0) | 0 (0·0) | 0 (0·0) | 0 (0·0) | 0 (0·0) | 0 (0·0) | 0 (0·0) | 0 (0·0) | 0 (0·0) |
| Priming vaccine | 0 (0·0) | 0 (0·0) | 0 (0·0) | 0 (0·0) | 0 (0·0) | 0 (0·0) | 0 (0·0) | 0 (0·0) | 0 (0·0) |
| Date dose 1 received | 0 (0·0) | 0 (0·0) | 0 (0·0) | 0 (0·0) | 0 (0·0) | 0 (0·0) | 0 (0·0) | 0 (0·0) | 0 (0·0) |
| Date dose 2 received | 0 (0·0) | 0 (0·0) | 0 (0·0) | 0 (0·0) | 0 (0·0) | 0 (0·0) | 0 (0·0) | 0 (0·0) | 0 (0·0) |
| Date dose 3 received | 0 (0·0) | 0 (0·0) | 0 (0·0) | 0 (0·0) | 0 (0·0) | 0 (0·0) | 0 (0·0) | 0 (0·0) | 0 (0·0) |
| Study day of blood draw | | | | | | | | | |
| Baseline | 1 (0·4) | 1 (0·8) | 0 (0·0) | 0 (0·0) | 0 (0·0) | 1 (1·7) | 0 (0·0) | 0 (0·0) | 0 (0·0) |
| Day 28 | 6 (2·3) | 2 (1·6) | 4 (3·1) | 0 (0·0) | 2 (3·9) | 1 (1·7) | 1 (1·7) | 1 (5·3) | 1 (5·3) |
| Six months | 14 (5·5) | 7 (5·5) | 7 (5·5) | 4 (8·0) | 4 (7·8) | 2 (3·4) | 2 (3·4) | 1 (5·3) | 1 (5·3) |
| 12 months | 15 (5·9) | 7 (5·5) | 8 (6·3) | 4 (8·0) | 3 (5·9) | 2 (3·4) | 4 (6·9) | 1 (5·3) | 1 (5·3) |
| 18 months | 35 (13·7) | 18 (14·1) | 17 (13·3) | 8 (16·0) | 6 (11·8) | 8 (13·6) | 10 (17·2) | 2 (10·5) | 1 (5·3) |
| 24 months | 34 (13·3) | 15 (11·7) | 19 (14·8) | 6 (12·0) | 6 (11·8) | 8 (13·6) | 10 (17·2) | 1 (5·3) | 3 (15·8) |

**Abbreviations:** CMI – cell-mediated immunity; IFN-γ – interferon-gamma; N – total number of participants; n – number with missing data.

**Footnotes:** [a] Ag1 - $CD4^+$ epitopes from the S1 subunit of the SARS-CoV-2 spike protein; [b] Ag2 - $CD4^+$ and $CD8^+$ epitopes from the S1 and S2 subunits of the SARS-CoV-2 spike protein; [c] Covariate completeness at baseline (day 0) is shown for age group, priming vaccine, and dates of doses 1–3; [c] Data are presented as n/N (%), unless otherwise indicated.

### *Missingness of 24 month IFN-γ Ag1 and Ag2 by baseline characteristics in the cell-mediated immunity sub-study*

We examined whether 24 month IFN-γ concentrations (Ag1 and Ag2 had identical counts) were differentially missing across baseline characteristics within the CMI sub-study (Supplementary Table 7). Missingness was slightly higher among participants aged 18–<50 years (13·9%, 25/180) than ≥50 years (11·8%, 9/76) and was similar by sex (13·7%, 18/131 in males; 12·8%, 16/125 in females). Differences across comorbidity groups were small in absolute terms and showed no consistent pattern. Continuous baseline variables for those missing IFN-γ concentration data (BMI and vaccine dosing intervals) were broadly similar across strata.

*Supplementary Table 7 Missingness of IFN-γ concentrations for Ag1[a] and Ag2[b] at 24 months in the cell-mediated immunity sub-study by baseline characteristics, overall, by study arm, and priming vaccine[c]*

| Variable | All Priming Strata | | | Primed with ChAdOx1-S | | Primed with BBIBP-CorV | | Primed with Gam-COVID-Vac | |
|---|---|---|---|---|---|---|---|---|---|
| | Total (N = 256) | Standard (N = 128) | Fractional (N= 128) | Standard (N = 50) | Fractional (N = 51) | Standard (N = 59) | Fractional (N = 58) | Standard (N = 19) | Fractional (N = 19) |
| | n/N (%) | n/N (%) | n/N (%) | n/N (%) | n/N (%) | n/N (%) | n/N (%) | n/N (%) | n/N (%) |
| 18–<50 years of age | 25/180 (13·9) | 12/90 (13·3) | 13/90 (14·4) | 6/42 (14·3) | 4/42 (9·5) | 6/35 (17·1) | 7/35 (20·0) | 0/13 (0·0) | 3/26 (11·5) |
| ≥50 years of age | 9/76 (11·8) | 3/38 (7·9) | 6/38 (15·8) | 0/8 (0·0) | 2/9 (22·2) | 2/24 (8·3) | 3/23 (13·0) | 1/6 (16·7) | 1/10 (10·0) |
| Male sex | 18/131 (13·7) | 5/64 (7·8) | 13/67 (19·4) | 4/28 (14·3) | 3/26 (11·5) | 1/26 (3·9) | 7/27 (25·9) | 0/10 (0·0) | 4/22 (18·2) |
| Female sex | 16/125 (12·8) | 10/64 (15·6) | 6/61 (9·8) | 2/22 (9·1) | 3/25 (12·0) | 7/33 (21·2) | 3/31 (9·7) | 1/9 (11·1) | 0/14 (0·0) |
| BMI, kg/m$^2$, median (IQR) | 23·9 (21·5-25·4) | 23·9 (21·4-26·7) | 24·7 (21·5-25·4) | 27·5 (23·9-31·1) | 23·4 (21·5-24·8) | 21·4 (20·6-22·8) | 22·6 (21·5-25·3) | 26·7 (26·7-26·7) | 27·4 (25·2-32·1) |
| Days between 1st and 2nd doses, median (IQR) | 38·5 (24-44) | 39 (26-49) | 34 (23-44) | 42 (41-53) | 43·5 (38-48) | 27 (22·5-33·5) | 29 (23-37) | 49 (49-49) | 60 (38·5-72) |
| Days between 2nd and study (3rd) dose, median (IQR) | 417 (391-494) | 400 (374-505) | 427 (403-479) | 505·5 (498-528) | 483·5 (404-523) | 376 (357·5-391) | 417 (400-430) | 414 (414-414) | 339 (407·5-455·5) |
| Reaction following 1st or 2nd dose | 2/44 (4·6) | 2/23 (8·7) | 0/21 (0·0) | 2/15 (13·3) | 0/12 (0·0) | 0/4 (0·0) | 0/6 (0·0) | 0/4 (0·0) | 1/6 (16·7) |
| Pain or fever medication for reaction | 0/11 (0·0) | 0/5 (0·0) | 0/6 (0·0) | 0/5 (0·0) | 0/5 (0·0) | NA | 0/1 (0·0) | NA | NA |
| Medical advice sought for reaction | 0/2 (0·0) | 0/1 (0·0) | 0/1 (0·0) | 0/1 (0·0) | 0/1 (0·0) | NA | NA | NA | NA |
| Symptoms of reaction resolved | 2/27 (7·4) | 2/17 (11·8) | 0/10 (0·0) | 2/10 (20·0) | 0/4 (0·0) | 0/3 (0·0) | 0/4 (0·0) | 0/4 (0·0) | 1/5 (20·0) |
| Self-reported prior SARS-CoV-2 | 17/147 (11·6) | 8/73 (11·0) | 9/74 (12·6) | 6/32 (18·8) | 2/35 (5·7) | 2/29 (6·9) | 4/25 (16·0) | 0/12 (0·0) | 4/29 (13·8) |
| Comorbidities | | | | | | | | | |
| Obesity (BMI ≥30 kg/m$^2$) | 2/52 (3·9) | 2/24 (8·3) | 0/28 (0·0) | 2/11 (18·2) | 0/12 (0·0) | 0/10 (0·0) | 0/12 (0·0) | 0/3 (0·0) | 0/9 (0·0) |
| Diabetes mellitus | 0/10 (0·0) | 0/8 (0·0) | 0/2 (0·0) | 0/2 (0·0) | 0/1 (0·0) | 0/4 (0·0) | 0/0 (0·0) | 0/2 (0·0) | 0/2 (0·0) |
| Cardiovascular disease | 0/20 (0·0) | 0/9 (0·0) | 0/11 (0·0) | 0/1 (0·0) | 0/5 (0·0) | 0/6 (0·0) | 0/5 (0·0) | 0/2 (0·0) | 0/2 (0·0) |
| Hypertension | 6/60 (10·0) | 3/27 (11·1) | 3/33 (9·1) | 2/10 (20·0) | 1/10 (10·0) | 1/14 (7·1) | 0/16 (0·0) | 0/3 (0·0) | 2/12 (16·7) |
| Cancer | NA | NA | NA | NA | NA | NA | NA | NA | NA |
| COPD | 0/4 (0·0) | 0/2 (0·0) | 0/2 (0·0) | NA | NA | 0/1 (0·0) | 0/1 (0·0) | 0/1 (0·0) | 0/1 (0·0) |
| Chronic kidney disease | 0/16 (0·0) | 0/7 (0·0) | 0/9 (0·0) | 0/3 (0·0) | 0/2 (0·0) | 0/4 (0·0) | 0/6 (0·0) | 0/3 (0·0) | 0/3 (0·0) |
| Chronic liver disease | 0/7 (0·0) | 0/2 (0·0) | 0/5 (0·0) | 0/1 (0·0) | NA | NA | 0/2 (0·0) | 0/1 (0·0) | 0/3 (0·0) |
| History of anaphylaxis | 0/3 (0·0) | 0/1 (0·0) | 0/2 (0·0) | 0/1 (0·0) | 0/2 (0·0) | NA | NA | NA | 0/1 (0·0) |
| Neurological disease | 0/4 (0·0) | 0/2 (0·0) | 0/2 (0·0) | 0/1 (0·0) | 0/1 (0·0) | 0/1 (0·0) | 0/1 (0·0) | NA | NA |
| On anticoagulant therapy | 0/11 (0·0) | 0/6 (0·0) | 0/5 (0·0) | 0/2 (0·0) | 0/1 (0·0) | 0/2 (0·0) | 0/3 (0·0) | 0/2 (0·0) | 0/3 (0·0) |
| Immunocompromised | NA | NA | NA | NA | NA | NA | NA | NA | NA |
| Mastocytosis causing recurrent anaphylaxis | NA | NA | NA | NA | NA | NA | NA | NA | NA |
| Cigarette user | 5/69 (7·3) | 2/34 (5·9) | 3/35 (8·6) | 1/15 (6·7) | 1/13 (7·7) | 0/10 (0·0) | 2/16 (12·5) | 1/9 (11·1) | 0/8 (0·0) |
| Currently pregnant | NA | NA | NA | NA | NA | NA | NA | NA | NA |

**Abbreviations**: BMI - body-mass index; IQR - interquartile range; IFN-γ – interferon-gamma; IQR – interquartile range; N – total number of participants in the stratum; n – number with missing data; NA – not applicable (no participants in that stratum). **Footnotes:** [a] Ag1 - $CD4^+$ epitopes from the S1 subunit of the SARS-CoV-2 spike protein; [b] Ag2 - $CD4^+$ and $CD8^+$ epitopes from the S1 and S2 subunits of the SARS-CoV-2 spike protein; [c] Data are presented as n/N (%), unless otherwise indicated

#### *IFN-γ Ag1 in the cell-mediated immunity sub-study*

Supplementary Table 8 summarises geometric mean IFN-γ Ag1 concentrations (IU/mL) for each study arm and the GMRs (fractional vs standard) at baseline, day 28, 6, 12, 18 and 24 months, reported overall and within priming strata (ChAdOx1-S, BBIBP-CorV, Gam-COVID-Vac), with estimates adjusted for age group, priming vaccine, dosing intervals and study day of blood draw (and additionally for baseline Ag1 at post-baseline visits).

*Supplementary Table 8 Geometric mean concentration of IFN-γ Ag1 and geometric mean ratio (fractional vs standard dose) at baseline, day 28, six, 12, 18, and 24 months, overall and stratified by priming vaccine*

| Priming Strata | GMC IFN-γ (IU/mL) (95% CI) | | GMR fractional/standard (95% CI) |
|---|---|---|---|
| | Standard Dose | Fractional Dose | |
| **Baseline[a]** | | | |
| All | 0·26 (0·20, 0·34) [n = 107] | 0·31 (0·23, 0·41)[n = 109] | 1·20 (0·83, 1·73); p = 0·339 |
| ChAdOx1-S | 0·37 (0·25, 0·56) [n = 46] | 0·38 (0·27, 0·54) [n =45 ] | 1·00 (0·59, 1·69); p = 0·993 |
| BBIBP-CorV | 0·16 (0·10, 0·26) [n = 44] | 0·20 (0·12, 0·34) [n =46 ] | 1·31 (0·68, 2·57); p = 0·414 |
| Gam-COVID-Vac | 0·33 (0·19, 0·58) [n = 17] | 0·53 (0·29, 0·98) [n = 18] | 1·77 (0·79, 4·00); p = 0·161 |
| **Day 28[b]** | | | |
| All | 0·46 (0·37, 0·58) [n = 113] | 0·60 (0·47, 0·77) [n = 112] | 1·19 (0·91, 1·56); p = 0·207 |
| ChAdOx1-S | 0·46 (0·32, 0·66) [n =48 ] | 0·55 (0·39, 0·80) [n = 49] | 1·15 (0·80, 1·65); p = 0·444 |
| BBIBP-CorV | 0·44 (0·31, 0·63) [n =48 ] | 0·68 (0·47, 0·99) [n = 49] | 1·35 (0·86, 2·14); p = 0·189 |
| Gam-COVID-Vac | 0·54 (0·30, 0·99) [n =17 ] | 0·53 (0·25, 1·20) [n = 17] | 0·94 (0·36, 2·41); p = 0·889 |
| ***Six months[b]*** | | | |
| All | 0·27 (0·21, 0·34) [n = 113] | 0·33 (0·26, 0·42) [n = 116] | 1·06 (0·77, 1·47); p = 0·719 |
| ChAdOx1-S | 0·20 (0·14, 0·30) [n = 42] | 0·26 (0·18, 0·39) [n =44 ] | 1·18 (0·75, 1·84); p = 0·475 |
| BBIBP-CorV | 0·31 (0·21, 0·45) [n = 53] | 0·38 (0·26, 0·54) [n = 55] | 0·95 (0·54, 1·69); p = 0·872 |
| Gam-COVID-Vac | 0·34 (0·18, 0·63) [n = 18] | 0·36 (0·18, 0·75) [n = 17] | 0·86 (0·38, 1·94); p = 0·701 |
| **12 months[b]** | | | |
| All | 0·28 (0·22, 0·36) [n = 92] | 0·30 (0·23, 0·39) [n = 93] | 0·98 (0·71, 1·37); p = 0·911 |
| ChAdOx1-S | 0·24 (0·15, 0·37) [n = 36] | 0·25 (0·16, 0·39) [n = 40] | 0·89 (0·54, 1·49); p = 0·659 |
| BBIBP-CorV | 0·30 (0·21, 0·44) [n = 40] | 0·40 (0·27, 0·59) [n = 39] | 1·39 (0·79, 2·44); p = 0·248 |
| Gam-COVID-Vac | 0·35 (0·22, 0·58) [n = 16] | 0·24 (0·14, 0·42) [n = 14] | 0·56 (0·30, 1·04); p = 0·065 |
| **18 months[b]** | | | |
| All | 0·21 (0·15, 0·28) [n = 83] | 0·28 (0·21, 0·37) [n = 91] | 1·18 (0·78, 1·78); p = 0·434 |
| ChAdOx1-S | 0·22 (0·14, 0·35) [n = 37] | 0·25 (0·17, 0·38) [n = 38] | 0·95 (0·56, 1·59); p = 0·833 |
| BBIBP-CorV | 0·18 (0·11, 0·31) [n = 32] | 0·31 (0·19, 0·52) [n = 41] | 1·76 (0·77, 3·99); p = 0·174 |
| Gam-COVID-Vac | 0·23 (0·11, 0·48) [n = 14] | 0·25 (0·14, 0·44) [n = 12] | 0·85 (0·34, 2·08); p = 0·701 |
| **24 months[b]** | | | |
| All | 0·27 (0·20, 0·37) [n = 113] | 0·35 (0·28, 0·46) [n = 109] | 1·17 (0·82, 1·66); p = 0·395 |
| ChAdOx1-S | 0·32 (0·19, 0·54) [n = 38] | 0·37 (0·26, 0·53) [n = 43] | 1·18 (0·77, 1·81); p = 0·447 |
| BBIBP-CorV | 0·24 (0·15, 0·39) [n = 46] | 0·35 (0·23, 0·53) [n = 45] | 1·23 (0·62, 2·44); p = 0·547 |
| Gam-COVID-Vac | 0·27 (0·15, 0·47) [n = 17] | 0·34 (0·18, 0·65) [n = 15] | 0·89 (0·44, 1·80); p = 0·734 |

**Abbreviations**: CI – confidence interval; GMR – geometric mean ratio; IFN-γ – interferon-gamma; IU/mL – international units per millilitre; n – number. **Footnotes:** [a] At baseline, GMRs are adjusted for age group, priming vaccine, days between doses 1–2, days between dose 2 and study dose, and study day of blood draw; [b] At day 28, six, 12, 18 and 24 months, GMRs are additionally adjusted for baseline IFN-γ (Ag1).

*IFN-γ Ag2 in the cell-mediated immunity sub-study*

Supplementary Table 9 summarises geometric mean IFN-γ Ag2 concentrations (IU/mL) for each study arm and the geometric mean ratios (GMRs; fractional vs standard) at baseline, day 28, six, 12, 18 and 24 months, reported overall and within priming strata (ChAdOx1-S, BBIBP-CorV, Gam-COVID-Vac).

*Supplementary Table 9 Geometric mean concentration of IFN-γ Ag2 and geometric mean ratio (fractional vs standard dose) at baseline, day 28, six, 12, 18, and 24 months, overall and stratified by priming vaccine.*

| Priming Strata | GMC IFN-γ (IU/mL) (95% CI) | | GMR fractional/standard (95% CI) |
|---|---|---|---|
| | Standard Dose | Fractional Dose | |
| **Baseline** | | | |
| All | 0·32 (0·24, 0·41) [n = 107] | 0·38 (0·30, 0·49) [n = 107] | 1·18 (0·83, 1·69) p = 0·362 |
| ChAdOx1-S | 0·43 (0·29, 0·64) [n = 48] | 0·43 (0·31, 0·61) [n = 47] | 1·01 (0·59, 1·73) p = 0·967 |
| BBIBP-CorV | 0·21 (0·14, 0·31) [n = 43] | 0·31 (0·19, 0·50) [n = 41] | 1·45 (0·79, 2·68) p = 0·230 |
| Gam-COVID-Vac | 0·39 (0·20, 0·76) [n = 16] | 0·44 (0·24, 0·80) [n = 19] | 1·13 (0·50, 2·53) p = 0·762 |
| **Day 28[a]** | | | |
| All | 0·64 (0·51, 0·79) [n = 113] | 0·75 (0·60, 0·94) [n = 109] | 1·06 (0·81, 1·38) p = 0·685 |
| ChAdOx1-S | 0·62 (0·45, 0·87) [n = 48] | 0·79 (0·56, 1·13) [n = 45] | 1·18 (0·81, 1·72) p = 0·383 |
| BBIBP-CorV | 0·65 (0·46, 0·93) [n = 48] | 0·76 (0·53, 1·10) [n = 48] | 0·97 (0·61, 1·55) p = 0·900 |
| Gam-COVID-Vac | 0·64 (0·35, 1·17) [n = 17] | 0·65 (0·40, 1·05) [n = 16] | 0·96 (0·49, 1·88) p = 0·897 |
| **Six months[b]** | | | |
| All | 0·34 (0·27, 0·42) [n = 116] | 0·42 (0·33, 0·54) [n = 109] | 1·14 (0·82, 1·59) p = 0·426 |
| ChAdOx1-S | 0·29 (0·20, 0·43) [n = 44] | 0·40 (0·28, 0·58) [n = 40] | 1·34 (0·87, 2·05) p = 0·183 |
| BBIBP-CorV | 0·38 (0·28, 0·52) [n = 55] | 0·42 (0·28, 0·62) [n = 53] | 0·95 (0·51, 1·76) p = 0·861 |
| Gam-COVID-Vac | 0·36 (0·21, 0·60) [n = 17] | 0·46 (0·23, 0·91) [n = 16] | 1·26 (0·61, 2·62) p = 0·515 |
| **12 months[b]** | | | |
| All | 0·44 (0·34, 0·57) [n = 104] | 0·59 (0·47, 0·73) [n = 106] | 1·14 (0·84, 1·54) p = 0·408 |
| ChAdOx1-S | 0·47 (0·31, 0·71) [n = 44] | 0·62 (0·43, 0·88) [n = 44] | 1·23 (0·78, 1·95) p = 0·367 |
| BBIBP-CorV | 0·40 (0·26, 0·61) [n = 45] | 0·58 (0·40, 0·84) [n = 46] | 1·32 (0·76, 2·33) p = 0·318 |
| Gam-COVID-Vac | 0·49 (0·28, 0·83) [n = 15] | 0·53 (0·33, 0·85) [n = 16] | 0·93 (0·61, 1·44) p = 0·745 |
| **18 months[b]** | | | |
| All | 0·42 (0·32, 0·55) [n = 92] | 0·49 (0·38, 0·64) [n = 104] | 0·99 (0·70, 1·40) p = 0·958 |
| ChAdOx1-S | 0·47 (0·30, 0·74) [n = 38] | 0·46 (0·30, 0·71) [n = 43] | 0·82 (0·51, 1·32) p = 0·412· |
| BBIBP-CorV | 0·40 (0·25, 0·62) [n = 41] | 0·54 (0·36, 0·81) [n = 43] | 1·30 (0·70, 2·41) p = 0·396 |
| Gam-COVID-Vac | 0·37 (0·21, 0·66) [n = 13] | 0·45 (0·24, 0·84) [n = 18] | 0·73 (0·35, 1·53) p = 0·387 |
| **24 months[b]** | | | |
| All | 0·34 (0·26, 0·45) [n = 105] | 0·42 (0·33, 0·54) [n = 106] | 1·06 (0·73, 1·54) p = 0·761 |
| ChAdOx1-S | 0·31 (0·19, 0·49) [n = 41] | 0·52 (0·36, 0·75) [n = 44] | 1·39 (0·85, 2·26) p = 0·182 |
| BBIBP-CorV | 0·36 (0·24, 0·56) [n = 46] | 0·34 (0·23, 0·50) [n = 46] | 0·71 (0·37, 1·36) p = 0·773 |
| Gam-COVID-Vac | 0·35 (0·19, 0·66) [n = 18] | 0·43 (0·21, 0·89) [n = 16] | 0·96 (0·38, 2·44) p = 0·927 |

**Abbreviations**: CI – confidence interval; GMR – geometric mean ratio; IFN-γ – interferon-gamma; IU/mL – international units per millilitre; n – number.

**Footnotes:** [a] At baseline, GMRs are adjusted for age group, priming vaccine, days between doses 1–2, days between dose 2 and study dose, and study day of blood draw; [b] At day 28, six, 12, 18 and 24 months, GMRs are additionally adjusted for baseline IFN-γ (Ag2).

Supplementary Table 10 presents IFN-γ responses (Ag1 and Ag2) stratified by age group and study arm across all visits. At baseline, concentrations were low across age groups, though point estimates were slightly higher in participants ≥50 years, particularly in the fractional-dose arm. By day 28, both age groups showed clear increases, with fractional-dose recipients tending to have higher GMCs than standard-dose recipients for both antigens. Responses were numerically higher in the ≥50 years group, though confidence intervals were wide.

By six and 12 months, responses had waned substantially in all groups, approaching baseline levels. In this period, GMCs were consistently lower in the <50 years group, whereas ≥50 years participants tended to sustain higher responses, especially for Ag2. Study-arm differences were small, with fractional-dose responses often equal to or slightly higher than standard-dose.

At 18 and 24 months, a divergence by age group was more evident. In participants <50 years, Ag1 responses remained low, while Ag2 rose modestly, particularly in the fractional-dose arm. In those ≥50 years, both Ag1 and Ag2 concentrations were higher than in younger adults, with fractional-dose recipients consistently showing the strongest responses. By 24 months, these differences persisted: responses in older adults were sustained at or above those in younger adults, and there was no indication of impaired durability with fractional dosing.

Overall, IFN-γ responses were transient, peaking at day 28 and declining thereafter, but older adults maintained higher concentrations at later time points than younger adults, particularly for Ag2, with no consistent disadvantage for fractional dosing.

*Supplementary Table 10 Geometric mean concentration of IFN-γ Ag1 and Ag2 at baseline, day 28, six, 12, 18, and 24 months, stratified by age group and study arm*

| Priming Strata | Ag1 | | Ag2 | |
|---|---|---|---|---|
| | GMC IFN-γ (IU/mL) (95% CI) | | GMC IFN-γ (IU/mL) (95% CI) | |
| | Standard Dose | Fractional Dose | Standard Dose | Fractional Dose |
| **Baseline** | | | | |
| 18–<50 years | 0·25 (0·18, 0·34) [n = 74] | 0·26 (0·19, 0·36) [n = 74] | 0·30 (0·22, 0·40) [n = 76] | 0·36 (0·27, 0·47) [n = 72] |
| ≥50 years | 0·29 (0·16, 0·51) [n = 33] | 0·43 (0·25, 0·74) [n = 35] | 0·37 (0·20, 0·67) [n = 31] | 0·44 (0·26, 0·76) [n = 35] |
| **Day 28** | | | | |
| 18–<50 years | 0·46 (0·35, 0·61) [n = 79] | 0·54 (0·41, 0·71) [n = 77] | 0·61 (0·47, 0·81) [n = 80] | 0·72 (0·55, 0·94) [n = 76] |
| ≥50 years | 0·47 (0·31, 0·71) [n = 34] | 0·77 (0·48, 1·23) [n = 35] | 0·70 (0·49, 0·99) [n = 33] | 0·84 (0·55, 1·28) [n = 33] |
| **Six months** | | | | |
| 18–<50 years | 0·24 (0·18, 0·31) [n = 76] | 0·30 (0·23, 0·40) [n = 81] | 0·29 (0·23, 0·38) [n = 82] | 0·40 (0·30, 0·53) [n = 77] |
| ≥50 years | 0·37 (0·22, 0·62) [n = 32] | 0·39 (0·22, 0·69) [n = 35] | 0·48 (0·31, 0·74) [n = 34] | 0·47 (0·28, 0·80) [n = 32] |
| **12 months** | | | | |
| 18–<50 years | 0·27 (0·20, 0·35) [n = 67] | 0·33 (0·24, 0·45) [n = 66] | 0·27 (0·20, 0·35) [n = 67] | 0·33 (0·24, 0·45) [n = 66] |
| ≥50 years | 0·34 (0·20, 0·56) [n = 25] | 0·25 (0·15, 0·41) [n = 27] | 0·34 (0·20, 0·56) [n = 25] | 0·25 (0·15, 0·41) [n = 27] |
| **18 months** | | | | |
| 18–<50 years | 0·19 (0·13, 0·27) [n = 60] | 0·22 (0·16, 0·31) [n = 63] | 0·40 (0·28, 0·56) [n = 65] | 0·45 (0·33, 0·60) [n = 73] |
| ≥50 years | 0·27 (0·15, 0·48) [n = 23] | 0·46 (0·29, 0·74) [n = 28] | 0·47 (0·29, 0·77) [n = 27] | 0·61 (0·35, 1·06) [n = 31] |
| **24 months** | | | | |
| 18–<50 years | 0·27 (0·19, 0·39) [n = 70] | 0·32 (0·24, 0·43) [n = 71] | 0·33 (0·23, 0·46) [n = 73] | 0·38 (0·28, 0·52) [n = 74] |
| ≥50 years | 0·28 (0·15, 0·50) [n = 31] | 0·43 (0·27, 0·71) [n = 32] | 0·36 (0·22, 0·60) [n = 32] | 0·52 (0·33, 0·81) [n = 32] |

**Abbreviations:** Ag – antigen; CI – confidence interval; GMC – geometric mean concentration; IFN-γ – interferon-gamma; IU/mL – international units per millilitre; n – number of participants with data

#### *Wuhan-Hu-1 SARS-CoV-2 sVNT inhibition*

Supplementary Table 11 summarises the median percentage inhibition of RBD–hACE2, a surrogate marker for the neutralising capacity against the original SARS-CoV-2 strain (Wuhan-Hu-1) overall and by priming group, stratified by study arm (standard and fractional doses). Baseline and day 28 results are included for completeness.

*Supplementary Table 11 Median Wuhan-Hu-1 SARS-CoV-2 sVNT percentage inhibition at baseline, day 28, six, 12, 18, and 24 months, stratified by study arm and priming vaccine*

| Priming Strata | Median inhibition % (IQR) | |
|---|---|---|
| | Standard Dose | Fractional Dose |
| **Baseline** | | |
| All | 81 (76–85) [n = 299] | 81 (77–84) [n = 300] |
| ChAdOx1-S | 81 (78–84) [n = 65] | 81 (78–85) [n = 64] |
| BBIBP-CorV | 80 (76–85) [n = 200] | 81 (77–84) [n = 200] |
| Gam-COVID-Vac | 80 (78–84) [n = 34] | 82 (79–84) [n = 36] |
| **day 28** | | |
| All | 81 (78–84) [n = 292] | 81 (78–84) [n = 295] |
| ChAdOx1-S | 81 (79–84) [n = 65] | 81 (77–84) [n = 62] |
| BBIBP-CorV | 81 (77–84) [n = 194] | 81 (78–83) [n = 198] |
| Gam-COVID-Vac | 80 (76–84) [n = 33] | 81 (79–84) [n = 35] |
| **Six months** | | |
| All | 89 (88–91) [n = 284] | 89 (88–91) [n = 290] |
| ChAdOx1-S | 89 (88–91) [n = 60] | 89 (86–90) [n = 60] |
| BBIBP-CorV | 89 (88–91) [n = 192] | 89 (88–91) [n = 195] |
| Gam-COVID-Vac | 89 (88–91) [n = 32] | 89 (88–91) [n = 35] |
| **12 months** | | |
| All | 89 (88–90) [n = 282] | 89 (87–90) [n = 285] |
| ChAdOx1-S | 89 (88–90) [n = 61] | 89 (88–90) [n = 61] |
| BBIBP-CorV | 89 (88–91) [n = 190] | 89 (87–90) [n = 190] |
| Gam-COVID-Vac | 89 (88–90) [n = 31] | 89 (88–90) [n = 34] |
| **18 months** | | |
| All | 88 (87–90) [n = 264] | 88 (86–90) [n = 265] |
| ChAdOx1-S | 88 (87–90) [n = 56] | 88 (87–90) [n = 58] |
| BBIBP-CorV | 88 (86–90) [n = 178] | 88 (86–90) [n = 173] |
| Gam-COVID-Vac | 89 (87–90) [n = 30] | 89 (87–91) [n = 34] |
| **24 months** | | |
| All | 88 (86–89) [n = 260] | 88 (86–90) [n = 260] |
| ChAdOx1-S | 88 (87–90) [n = 58] | 89 (87–90) [n = 58] |
| BBIBP-CorV | 87 (84–89) [n = 172] | 88 (85–90) [n = 170] |
| Gam-COVID-Vac | 88 (87–89) [n = 30] | 89 (87–91) [n = 32] |

**Abbreviations**: IQR – interquartile range

#### *Omicron BA.1 SARS-CoV-2 sVNT inhibition*

Supplementary Table 12 displays the median RBD–hACE2 percentage inhibition against the SARS-CoV-2 Omicron BA.1 variant at six and 12 months post-booster by priming vaccine and study arm. Baseline and day 28 results are included for completeness.

*Supplementary Table 12 Median Omicron BA.1 SARS-CoV-2 RBD–hACE2 percentage inhibition at baseline, day 28, six, 12, 18, and 24 months, stratified by study arm and priming vaccine*

| Priming Strata | Median inhibition % (IQR) | |
|---|---|---|
| | Standard Dose | Fractional Dose |
| Baseline | | |
| All | 52 (17–77) [n = 299] | 51 (18–76) [n = 300] |
| ChAdOx1-S | 69 (38–81) [n = 65] | 59 (37–75) [n = 64] |
| BBIBP-CorV | 42 (7–72) [n = 200] | 43 (13–72) [n = 200] |
| Gam-COVID-Vac | 61 (36–76) [n = 34] | 68 (41–81) [n = 36] |
| Day 28 | | |
| All | 82 (75–85) [n = 292] | 81 (75–84) [n = 295] |
| ChAdOx1-S | 82 (80–84) [n = 65] | 80 (77–83) [n = 62] |
| BBIBP-CorV | 80 (72–85) [n = 194] | 81 (72–84) [n = 198] |
| Gam-COVID-Vac | 83 (78–85) [n = 33] | 82 (76–85) [n = 35] |
| Six months | | |
| All | 74 (46–87) [n = 284] | 77 (48–87) [n = 290] |
| ChAdOx1-S | 80 (60–86) [n = 60] | 78 (55–86) [n = 60] |
| BBIBP-CorV | 73 (40–86) [n = 192] | 76 (44–88) [n = 195] |
| Gam-COVID-Vac | 69 (43–88) [n = 32] | 81 (60–88) [n = 35] |
| 12 months | | |
| All | 76 (44–87) [n = 282] | 79 (40–87) [n = 285] |
| ChAdOx1-S | 82 (65–88) [n = 61] | 74 (50–86) [n = 61] |
| BBIBP-CorV | 72 (35–87) [n = 190] | 79 (37–87) [n = 190] |
| Gam-COVID-Vac | 79 (49–88) [n = 31] | 84 (62–89) [n = 34] |
| 18 months | | |
| All | 81 (61–86) [n = 264] | 81 (64–86) [n = 265] |
| ChAdOx1-S | 82 (72–85) [n = 56] | 81 (67–87) [n = 58] |
| BBIBP-CorV | 80 (59–86) [n = 178] | 81 (60–86) [n = 173] |
| Gam-COVID-Vac | 82 (75–85) [n = 30] | 84 (76–87) [n = 34] |
| 24 months | | |
| All | 84 (71–88) [n = 260] | 85 (70–88) [n = 260] |
| ChAdOx1-S | 85 (78– 88) [n = 58] | 85 (81–88) [n = 58] |
| BBIBP-CorV | 84 (65–87) [n = 172] | 84 (66–87) [n = 170] |
| Gam-COVID-Vac | 85 (71–87) [n = 30] | 87 (82–89) [n = 32] |

**Abbreviations**: IQR – interquartile range

#### *Documented and Suspected Intercurrent SARS-CoV-2 Infections*

Documented SARS-CoV-2 infections are shown in Supplementary Figure 1, with a cumulative incidence of 4·7% (28/601) by 24 months.

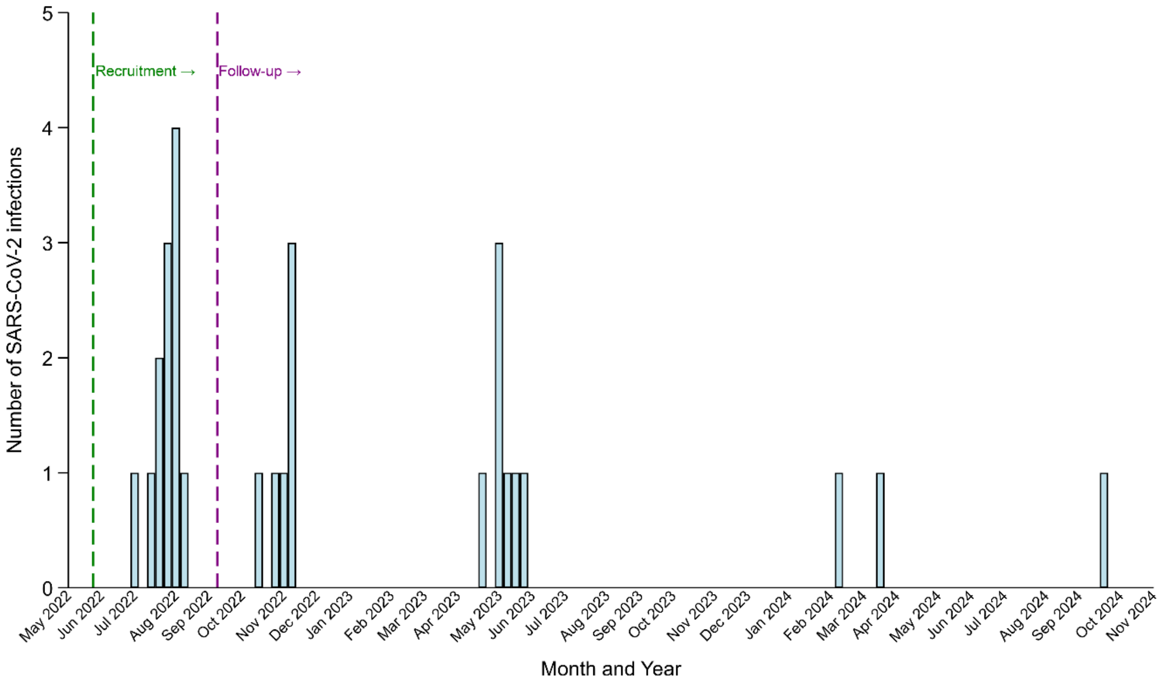


*Supplementary Figure 1 SARS-CoV-2 infections by study week from trial baseline to 24 month visit window (n = 28)*

### Adverse and Serious Adverse Events

From baseline to 24 months post-booster, 76 AEs and 53 SAEs were recorded (Supplementary Tables 13 and 14, and Supplementary Figure 2).

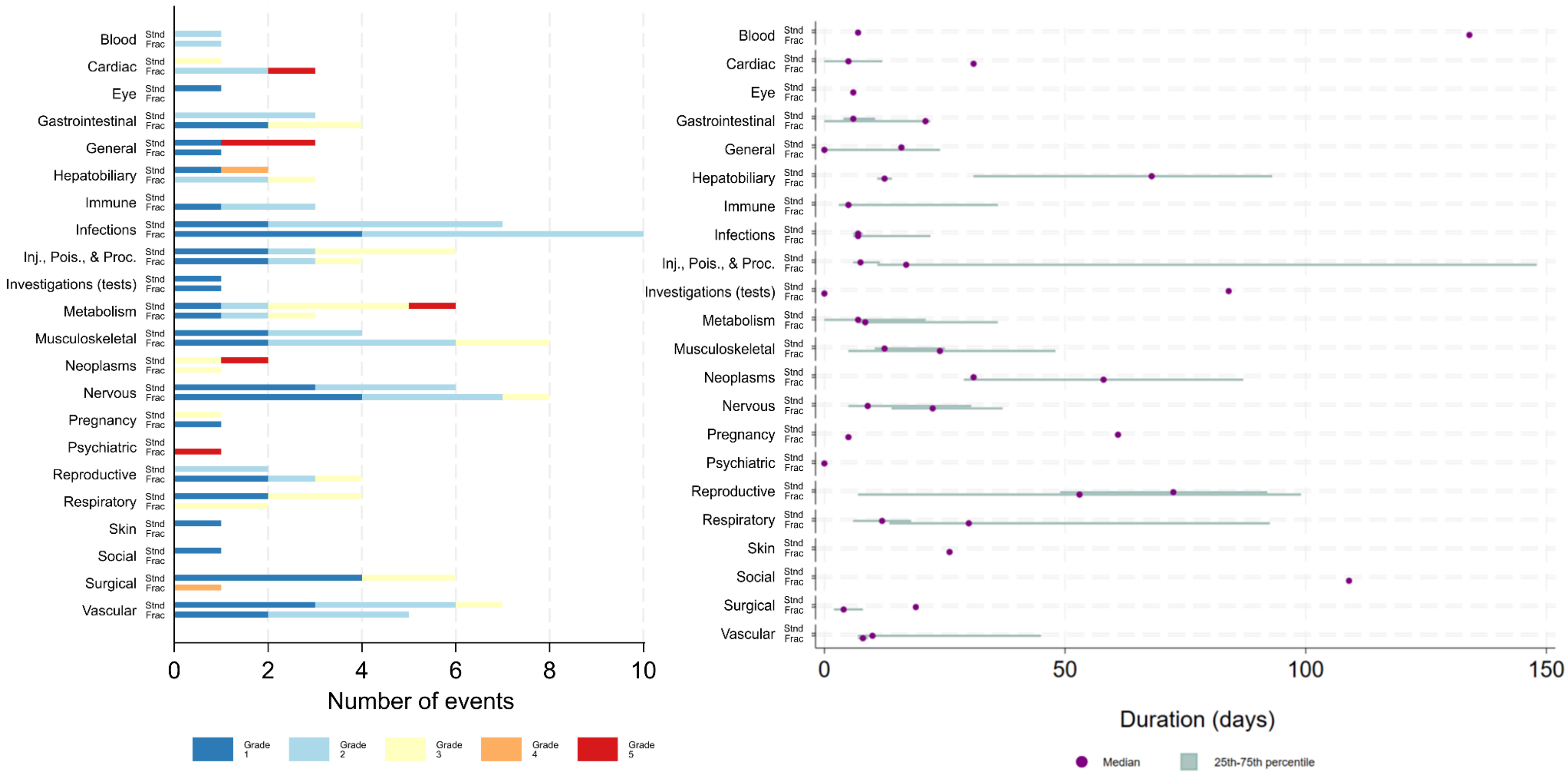


*Supplementary Figure 2 Severity and duration of adverse events by MedDRA System Organ Class (SOC) and study arm. Left panel: number and severity (Grade 1 – 5) of adverse events reported in each MedDRA SOC, stratified by study arm. Right panel: Median duration (diamond) and interquartile range (horizontal bars) of adverse events by SOC and study arm.* ***SOC abbreviations****: Blood - Blood and lymphatic system disorders; Cardiac - Cardiac disorders; Eye - Eye disorders; Gastrointestinal - Gastrointestinal disorders; General - General disorders and administration site conditions; Hepatobiliary - Hepatobiliary disorders; Immune - Immune system disorders; Infections - Infections and infestations; Inj., Pois., & Proc. - Injury, poisoning and procedural complications; Investigations - Investigations (e.g. abnormal lab tests or vitals); Metabolism - Metabolism and nutrition disorders; Musculoskeletal - Musculoskeletal and connective tissue disorders; Neoplasms - Neoplasms benign, malignant and unspecified; Nervous - Nervous system disorders; Pregnancy - Pregnancy, puerperium and perinatal conditions; Psychiatric - Psychiatric disorders; Reproductive - Reproductive system and breast disorders; Respiratory - Respiratory, thoracic and mediastinal disorders; Skin - Skin and subcutaneous tissue disorders; Social - Social circumstances; Surgical - Surgical and medical procedures; Vascular - Vascular disorders.*

Supplementary Table 13 provides an overview of adverse events (AEs) reported among participants, classified by their relationship to the study vaccine, severity, and timelines.

*Supplementary Table 13 Adverse events (n=76), classified by study arm, MedDRA term, relationship to study vaccine, severity, onset relative to vaccination, duration, and outcome*

| Case number | Study arm | MedDRA term | Relationship to Study Vaccine [a] | Severity [b] | Administration of study vaccine to AE start date (days) | Duration of event (days) | Outcome |
|---|---|---|---|---|---|---|---|
| 1 | Standard | Abdominal cramps | Unrelated | Moderate | 3 | 0 | Resolved |
| 2 | Standard | Arthralgia | Unrelated | Moderate | 1 | 40 | Resolved |
| 3 | Standard | Back pain | Unrelated | Mild | 175 | 2 | Resolved |
| 4 | Standard | Back pain | Unrelated | Mild | 21 | 56 | Resolved |
| 5 | Standard | Blood glucose increased | Unrelated | Mild | 55 | 0 | Resolved |
| 6 | Standard | Chronic bronchitis | Unrelated | Mild | 173 | 40 | Resolved |
| 7 | Standard | Dyspnoea | Unrelated | Mild | 6 | 20 | Resolved with sequelae |
| 8 | Standard | Eczema | Possible | Mild | 17 | 26 | Resolved |
| 9 | Standard | Eye inflammation | Unrelated | Mild | 64 | 6 | Resolved |
| 10 | Standard | Fatigue | Possible | Mild | 2 | 24 | Resolved |
| 11 | Standard | Furunculosis | Possible | Moderate | 33 | 62 | Resolved |
| 12 | Standard | Gallbladder disorder | Unrelated | Mild | 94 | 14 | Resolved |
| 13 | Standard | Gallstones | Unrelated | Mild | 174 | 7 | Resolved |
| 14 | Standard | Headache | Possible | Mild | 16 | 69 | Resolved |
| 15 | Standard | Headache | Possible | Mild | 0 | 27 | Resolved |
| 16 | Standard | Headache | Possible | Mild | 8 | 18 | Resolved |
| 17 | Standard | Headache | Possible | Moderate | 8 | 14 | Resolved |
| 18 | Standard | Headache | Possible | Moderate | 48 | 12 | Resolved |
| 19 | Standard | Headache | Unrelated | Moderate | 0 | 37 | Resolved with sequelae |
| 20 | Standard | Hypertension | Unrelated | Mild | 25 | 3 | Resolved |
| 21 | Standard | Hypertension | Unrelated | Mild | 328 | 8 | Resolved |
| 22 | Standard | Hypertension | Possible | Mild | 7 | 8 | Resolved |
| 23 | Standard | Hypertension | Possible | Moderate | 83 | 10 | Resolved with sequelae |
| 24 | Standard | Hypertension | Possible | Moderate | 10 | 9 | Resolved with sequelae |
| 25 | Standard | Menstruation irregular | Possible | Moderate | 3 | 99 | Resolved |
| 26 | Standard | Natural menopause | Possible | Mild | 0 | 109 | Resolved |

| | | | | | | | |
|---|---|---|---|---|---|---|---|
| 27 | Standard | Radius fracture | Unrelated | Mild | 190 | 5 | Resolved |
| 28 | Standard | Upper respiratory tract infection | Unrelated | Mild | 26 | 7 | Resolved |
| 29 | Standard | Upper respiratory tract infection | Unrelated | Moderate | 1 | 6 | Resolved |
| 30 | Standard | Dental care | Unrelated | Mild | 56 | 21 | Resolved |
| 31 | Standard | Dental care | Unrelated | Mild | 69 | 0 | Resolved |
| 32 | Standard | Dental care | Unrelated | Mild | 84 | 2 | Resolved |
| 33 | Standard | Dental care | Unrelated | Mild | 91 | 8 | Resolved |
| 34 | Standard | Eye injury | Unrelated | Moderate | 76 | 11 | Resolved |
| 35 | Standard | Limb injury | Unrelated | Mild | 106 | 21 | Resolved |
| 36 | Standard | Acute upper respiratory tract infection | Unrelated | Mild | 22 | 5 | Resolved |
| 37 | Standard | Acute respiratory tract infection | Unrelated | Moderate | 10 | 22 | Resolved |
| 38 | Standard | Blood loss anaemia | Unrelated | Moderate | 65 | 134 | Resolved |
| 39 | Fractional | Amenorrhea | Possible | Mild | 45 | 70 | Resolved |
| 40 | Fractional | Anaemia | Unrelated | Moderate | 20 | 7 | Resolved |
| 41 | Fractional | Arthralgia | Unrelated | Moderate | 1 | 10 | Resolved |
| 42 | Fractional | Back injury | Unrelated | Moderate | 132 | 7 | Resolved with sequelae |
| 43 | Fractional | Back pain | Unrelated | Moderate | 8 | 12 | Resolved |
| 44 | Fractional | Chest discomfort | Unrelated | Mild | 11 | 16 | Resolved with sequelae |
| 45 | Fractional | Chest pain | Possible | Moderate | 44 | 5 | Resolved |
| 46 | Fractional | Chronic gastritis | Unrelated | Mild | 56 | 14 | Resolved |
| 47 | Fractional | Dental caries | Unrelated | Mild | 144 | 3 | Resolved |
| 48 | Fractional | Diabetes mellitus | Unrelated | Mild | 79 | 0 | Resolved |
| 49 | Fractional | Furuncle | Possible | Mild | 12 | 8 | Resolved |
| 50 | Fractional | Furuncle | Possible | Mild | 27 | 8 | Resolved |
| 51 | Fractional | Headache | Possible | Mild | 1 | 30 | Resolved |
| 52 | Fractional | Headache | Possible | Mild | 23 | 4 | Resolved |
| 53 | Fractional | Headache | Possible | Mild | 8 | 6 | Resolved |
| 54 | Fractional | Headaches | Unrelated | Moderate | 143 | 31 | Resolved |
| 55 | Fractional | Heart rate irregular | Possible | Mild | 31 | 84 | Resolved with sequelae |
| 56 | Fractional | Hypersensitivity | Unrelated | Moderate | 67 | 3 | Resolved |
| 57 | Fractional | Hypertension | Possible | Mild | 8 | 97 | Resolved |
| 58 | Fractional | Hypertension | Possible | Mild | 39 | 45 | Resolved with sequelae |
| 59 | Fractional | Hypertension | Unrelated | Moderate | 8 | 7 | Resolved |

| | | | | | | | |
|---|---|---|---|---|---|---|---|
| 60 | Fractional | Irregular menstruation | Possible | Mild | 8 | 109 | Resolved |
| 61 | Fractional | Menstruation irregular | Possible | Moderate | 39 | 75 | Resolved |
| 62 | Fractional | Myalgia | Unrelated | Mild | 64 | 44 | Resolved |
| 63 | Fractional | Palpitations | Possible | Moderate | 3 | 12 | Resolved |
| 64 | Fractional | Pregnancy | Unrelated | Mild | 116 | 61 | Resolved |
| 65 | Fractional | Soft tissue injury | Unrelated | Mild | 163 | 15 | Resolved |
| 66 | Fractional | Syncope | Unrelated | Mild | 37 | 0 | Resolved |
| 67 | Fractional | Upper respiratory tract infection | Unrelated | Moderate | 2 | 27 | Resolved |
| 68 | Fractional | Varicella | Unrelated | Moderate | 344 | 7 | Resolved |
| 69 | Fractional | Tooth infection | Unrelated | Moderate | 16 | 7 | Resolved |
| 70 | Fractional | Face injury | Unrelated | Mild | 86 | 8 | Resolved |
| 71 | Fractional | Lower limb fracture | Unrelated | Mild | 103 | 20 | Resolved |
| 72 | Fractional | Allergy to chemicals | Unrelated | Moderate | 24 | 5 | Resolved |
| 73 | Fractional | Acute upper respiratory tract infection | Unrelated | Mild | 21 | 5 | Resolved |
| 74 | Fractional | Acute respiratory tract infection | Unrelated | Mild | 23 | 5 | Resolved |
| 75 | Fractional | Acute respiratory tract infection | Unrelated | Moderate | 18 | 6 | Resolved |
| 76 | Fractional | Dust allergy | Possible | Mild | 70 | 36 | Resolved |

**Abbreviations:** AE – adverse event; MedDRA – Medical Dictionary for Regulatory Activities. **Footnotes:** [a] Relationship to study vaccine was categorised as unrelated, possible, probable, or definite. [b] Severity was graded as mild (grade 1), moderate (grade 2), severe (grade 3), potentially life-threatening (grade 4), or fatal (grade 5).

### Serious Adverse Events

Supplementary Table 14 shows all serious adverse events recorded within the 24 month visit window.

*Supplementary Table 14 Serious adverse events (n=53), classified by study arm, MedDRA term, relationship to study vaccine, severity, onset relative to vaccination, duration, and outcome*

| Case number | Study arm | MedDRA term | Relationship to Study vaccine [a] | Severity [b] | Administration of study vaccine to AE start date (days) | Duration of event (days) | Outcome |
|---|---|---|---|---|---|---|---|
| 1 | Standard | Back pain | Unrelated | Moderate | 38 | 8 | Resolved |
| 2 | Standard | Cholecystectomy | Unrelated | Severe | 118 | 5 | Resolved |
| 3 | Standard | Cholecystectomy | Unrelated | Severe | 88 | 3 | Resolved |
| 4 | Standard | Cholecystitis | Unrelated | Potentially life-threatening | 11 | 11 | Resolved with sequelae |
| 5 | Standard | Chronic bronchitis | Unrelated | Severe | 35 | 7 | Resolved with sequelae |
| 6 | Standard | Chronic obstructive pulmonary disease | Unrelated | Severe | 485 | 145 | Resolved with sequelae |

| | | | | | | | |
|---|---|---|---|---|---|---|---|
| 7 | Standard | Colorectal cancer stage II | Unrelated | Severe | 293 | 29 | Resolved with sequelae |
| 8 | Standard | Coronary artery stenosis | Unrelated | Severe | 477 | 31 | Resolved with sequelae |
| 9 | Standard | Cystocele | Unrelated | Moderate | 177 | 7 | Resolved |
| 10 | Standard | Diabetes mellitus | Unrelated | Moderate | 294 | 7 | Resolved with sequelae |
| 11 | Standard | Diabetes mellitus | Unrelated | Severe | 17 | 36 | Resolved with sequelae |
| 12 | Standard | Diabetes mellitus | Unrelated | Severe | 82 | 6 | Resolved with sequelae |
| 13 | Standard | Diabetes mellitus inadequate control | Unrelated | Severe | 45 | 10 | Resolved with sequelae |
| 14 | Standard | Ectopic pregnancy | Unrelated | Severe | 130 | 5 | Resolved |
| 15 | Standard | Gastric cancer | Unrelated | Fatal | 44 | 87 | Fatal† |
| 16 | Standard | Gastric polyps | Unrelated | Moderate | 475 | 22 | Resolved |
| 17 | Standard | Hypertension | Unrelated | Moderate | 46 | 7 | Resolved with sequelae |
| 18 | Standard | Hypertension | Unrelated | Severe | 494 | 7 | Resolved with sequelae |
| 19 | Standard | Pyelonephritis chronic | Unrelated | Moderate | 385 | 7 | Resolved |
| 20 | Standard | Respiratory infection | Unrelated | Moderate | 99 | 7 | Resolved |
| 21 | Standard | Sudden death | Unrelated | Fatal | 480 | 0 | Fatal† |
| 22 | Standard | Sudden death | Unrelated | Fatal | 518 | 0 | Fatal† |
| 23 | Standard | Large intestine polyp | Unrelated | Moderate | 540 | 21 | Resolved |
| 24 | Standard | Lower limb fracture | Unrelated | Severe | 1 | 13 | Resolved |
| 25 | Standard | Lower limb fracture | Unrelated | Severe | 64 | 273 | Resolved |
| 26 | Standard | Meniscus injury | Unrelated | Severe | 369 | 148 | Resolved |
| 27 | Standard | Decompensated diabetes | Unrelated | Fatal | 124 | 40 | Fatal† |
| 28 | Fractional | Arm amputation | Unrelated | Potentially life-threatening | 352 | 19 | Resolved with sequelae |
| 29 | Fractional | Asthma, unspecified type, with status asthmaticus | Unrelated | Severe | 154 | 6 | Resolved with sequelae |
| 30 | Fractional | Back injury | Unrelated | Severe | 84 | 5 | Resolved with sequelae |
| 31 | Fractional | Back pain | Unrelated | Moderate | 78 | 13 | Resolved |
| 32 | Fractional | Back pain | Unrelated | Severe | 160 | 7 | Resolved with sequelae |
| 33 | Fractional | Cerebrovascular accident | Unrelated | Severe | 322 | 11 | Resolved with sequelae |
| 34 | Fractional | Cholecystitis | Unrelated | Severe | 21 | 93 | Resolved |
| 35 | Fractional | Cholelithiasis | Unrelated | Moderate | 139 | 31 | Resolved |
| 36 | Fractional | Chronic pyelonephritis | Unrelated | Moderate | 265 | 7 | Resolved |
| 37 | Fractional | Completed suicide | Unrelated | Fatal | 263 | 0 | Fatal† |
| 38 | Fractional | Diabetes mellitus | Unrelated | Severe | 98 | 7 | Resolved with sequelae |

| | | | | | | | |
|---|---|---|---|---|---|---|---|
| 39 | Fractional | Gallbladder disorder | Unrelated | Moderate | 374 | 68 | Resolved |
| 40 | Fractional | Gout | Unrelated | Moderate | 11 | 21 | Resolved with sequelae |
| 41 | Fractional | Gouty arthritis | Unrelated | Severe | 51 | 11 | Resolved with sequelae |
| 42 | Fractional | Haemorrhoids | Unrelated | Severe | 323 | 5 | Resolved |
| 43 | Fractional | Headache | Unrelated | Moderate | 126 | 7 | Resolved with sequelae |
| 44 | Fractional | Hypertension | Unrelated | Moderate | 80 | 7 | Resolved |
| 45 | Fractional | Hypertension | Unrelated | Moderate | 292 | 10 | Resolved with sequelae |
| 46 | Fractional | Myocardial infarction | Unrelated | Fatal | 509 | 0 | Fatal† |
| 47 | Fractional | Pneumonia mycoplasmal | Unrelated | Severe | 166 | 18 | Resolved |
| 48 | Fractional | Pyelonephritis acute | Unrelated | Moderate | 543 | 10 | Resolved |
| 49 | Fractional | Uterine leiomyoma | Unrelated | Severe | 477 | 28 | Resolved |
| 50 | Fractional | Cerebral cyst | Unrelated | Moderate | 246 | 167 | Resolved |
| 51 | Fractional | Intervertebral disc disorder | Unrelated | Moderate | 690 | 30 | Resolved with sequelae |
| 52 | Fractional | Hepatic cancer | Unrelated | Severe | 513 | 31 | Resolved with sequelae |
| 53 | Fractional | Ulcerative gastritis | Unrelated | Severe | 153 | 7 | Resolved with sequelae |

**Abbreviations:** AE – adverse event; MedDRA – Medical Dictionary for Regulatory Activities. **Footnotes:** [a] Relationship to study vaccine was categorised as unrelated, possible, probable, or definite. [b] Severity was graded as mild (grade 1), moderate (grade 2), severe (grade 3), potentially life-threatening (grade 4), or fatal (grade 5).

### References


1. Rubin DB. Multiple imputation for survey nonresponse. New York: Wiley, 1987.
2. White IR, Royston P, Wood AM. Multiple imputation using chained equations: Issues and guidance for practice. *Stat Med* 2011; 30(4): 377-99.
